# Untargeted viral metagenomics of indoor air as a novel surveillance tool for respiratory, enteric and skin viruses

**DOI:** 10.1101/2024.10.11.24315306

**Authors:** Mustafa Karatas, Caspar Geenen, Els Keyaerts, Lore Budts, Joren Raymenants, Charlotte Eggers, Bastiaan Craessaerts, Emmanuel André, Jelle Matthijnssens

**Author notes:** These authors contributed equally (Emmanuel André, Jelle Matthijnssens).

## Abstract

Conventional hospital-based infectious disease surveillance struggles to detect mild or asymptomatic infections and incurs high costs for large-scale testing during outbreaks. In contrast, environmental surveillance can effectively monitor viral circulation and capture asymptomatic or mildly symptomatic infections that may otherwise go unnoticed. This longitudinal study explores the use of indoor air, in combination with targeted qPCR panels and untargeted viral metagenomics, as a novel virus surveillance tool. Indoor air samples were collected weekly from a daycare center in Leuven, Belgium, over a 12-month period using active indoor air sampling, followed by screening using respiratory and enteric qPCR panels, as well as untargeted viral metagenomics.

Human-associated viruses were detected in 95.2% (40/42) of samples, with MW polyomavirus being the most prevalent at 80.9%. Several other respiratory viruses (e.g., rhinoviruses, RSV-B) and enteric viruses (e.g.. rotavirus, astrovirus, adenovirus) were identified, correlating with their known epidemiological circulation patterns. Metagenomics resulted in multiple complete viral genome reconstructions, allowing discrimination of viral subtypes and the identification of coinfections with closely related virus variants. Finally, a plethora of animal, insect, fungal and plant-infecting viruses could be detected, representing the indoor and outdoor environment. Indoor air surveillance can be a robust complementary tool for virus surveillance.

## Introduction

Conventionally, infectious disease surveillance has revolved around patient-centered approaches, focusing on specific diseases or syndromes (e.g., respiratory, enteric symptoms) using methods such as pathogen culturing and molecular techniques (PCR or qPCR panels) on individual patient samples^1^. While valuable for immediate patient care, these methods often fail to detect novel viruses, can be invasive, and become costly when extensive testing is required^2^. In addition, consent is required, which could be challenging, especially for infections that cause many mild or even asymptomatic cases but can also lead to severe presentations. The COVID-19 pandemic underscored these challenges and highlighted the need for scalable surveillance methods. In response, environmental surveillance has emerged as a crucial alternative, allowing for the monitoring of viral circulation in communities without relying on direct patient sampling.

Over the past decade, environmental surveillance for pathogens, using media such as wastewater, soil, rivers, and indoor air, has attracted growing interest, with wastewater being the most widely utilized method^3–5^. Wastewater analysis has effectively complemented other surveillance systems by tracking SARS-CoV-2, its variants of concern, and other respiratory and enteric pathogens using targeted (quantitative) PCR methods, thereby reinforcing its importance alongside conventional patient-centered surveillance^6^. This approach offers insights into prevalence and sometimes genetic diversity of pathogens across broader populations and enables the early detection of emerging strains and trends^7,8^. However, wastewater surveillance has limitations, including reduced precision in pinpointing specific locations or population subgroups, potential delays or signal dilution due to rainfall, and dependency on water usage^9^. Recently, our teams and others have demonstrated the potential of active indoor air sampling for monitoring viruses^10–13^. This method has successfully identified a variety of viruses, including respiratory and enteric viruses (e.g., respiratory syncytial virus, influenza C, human parainfluenza virus) in community settings.

The methodologies employed to detect pathogens in environmental surveillance are usually similar to those used in clinical diagnostics^1,14^, often using molecular based single- or multi-targeted (qPCR or multiplex qPCR) tests. Targeted test methods tend to have high sensitivity and specificity. Previously, a number of studies has focused on the detection of viruses and bacteria with targeted methods from indoor air of various community settings^10,12,13^, revealing temporal trends of virus circulation among specific populations and highlighting the early warning potential of indoor air sampling in real-world situations.

However, for more precise genetic identification or subtyping, additional techniques such as Sanger sequencing or Whole Genome Sequencing (WGS) are often necessary. Characterizing novel or variant viruses can be challenging with both methods, because they require specific sequence knowledge of viruses of interest. However, Next Generation Sequencing (NGS)-based ’untargeted’ shotgun metagenomics can overcome this limitation. These methods often use a combination of viral particle purification, followed by deep sequencing using various NGS technologies and appropriate bio-informatics tools^15^. An established method, Novel Enrichment Techniques of Viromes (NetoVIR)^16^, has proven effective for viral enrichment across various sample types, including fecal^17,18^, insect^19,20^, and plant samples^21^, facilitating virome analysis with minimal bias and the discovery of novel viruses. This approach can identify the pathogen and provide genomic insights as a ’pathogen-agnostic’ tool^22^. For indoor air surveillance, NGS faces challenges such as low biomass and contaminating nucleic acid, despite its relatively cleaner nature compared to soil, wastewater or stool^23^. Nevertheless, *Minor et al.* used RNA virus-targeted next-generation sequencing (NGS) to analyze 20 samples from various indoor community settings. They detected respiratory viruses such as influenza A and C, but also rotaviruses and astrovirus, which are transmitted through the fecal-oral route^24^.

In this study, we employed weekly active air sampling at a daycare center for children aged 0-3 years, spanning a full year, and analyzed these samples using untargeted viral metagenomics. Our objective was to investigate the potential of air sampling for surveillance, by employing both reference-guided and de-novo bioinformatics approaches to achieve a better understanding of the indoor air virome and its dynamics.

## Results

### 1. Study design

Air samples were collected longitudinally from a daycare center unit in Leuven, Belgium, between January and December 2022. Particles suspended in indoor air were collected using an AerosolSense sampler (Thermo Fisher Scientific, Waltham, MA) for two hours. Ninety samples were collected from the same location, near the ventilation system’s air extraction point in the changing room of the daycare unit. All ninety samples were processed with a respiratory qPCR panel, while forty-two available samples were further processed using an enteric qPCR panel and viral metagenomics (*see Methods*). For the latter, samples were enriched for viral-like particles (VLPs) using the NetoVIR protocol and sequenced on the Illumina NovaSeq 6000 platform. On average, 25 million reads were produced per sample. Produced reads were then further analyzed using two bioinformatics approaches: *EsViritu,* referred to as “reference guided assembly”, and *ViPER,* referred to as “*de-novo* assembly” (*see methods*). Findings from these two approaches are explained and discussed together, unless specified otherwise.

### 2. Untargeted shotgun metagenomics revealed a spectrum of enteric, respiratory and skin viruses in indoor air

Using reference-guided assembly, we identified human-associated viruses in 40 out of 42 samples (95%, list of viruses identified in each sample with read counts can be found in **Supplementary information 1**). *De novo* assembly identified human-associated viruses in 29 out of 42 samples (69%) (**Figure 1. A. & B**.). Several respiratory viruses were detected throughout the year, including human bocavirus 1, 2 and 3 (n=18, species *Bocaparvovirus primate1-2*), *Enterovirus alpharhino* and *Enterovirus cerhino* (formerly *Rhinovirus A* (n=9) and *C* (n=5)), human adenovirus 6 (n=1), viruses belonging to the genera *Betacoronavirus* (Human coronavirus HKU1, n=3 and OC43, n=1) and respiratory syncytial virus B / RSV-B, n=2). WU polyomavirus was consistently detected from the end of May until the summer holiday closure in July, and it accounted for the highest total number of human-infecting virus reads in our study. Among the identified respiratory viruses, we obtained complete genomes of WU polyomavirus and human bocavirus 1.

**Figure 1.**
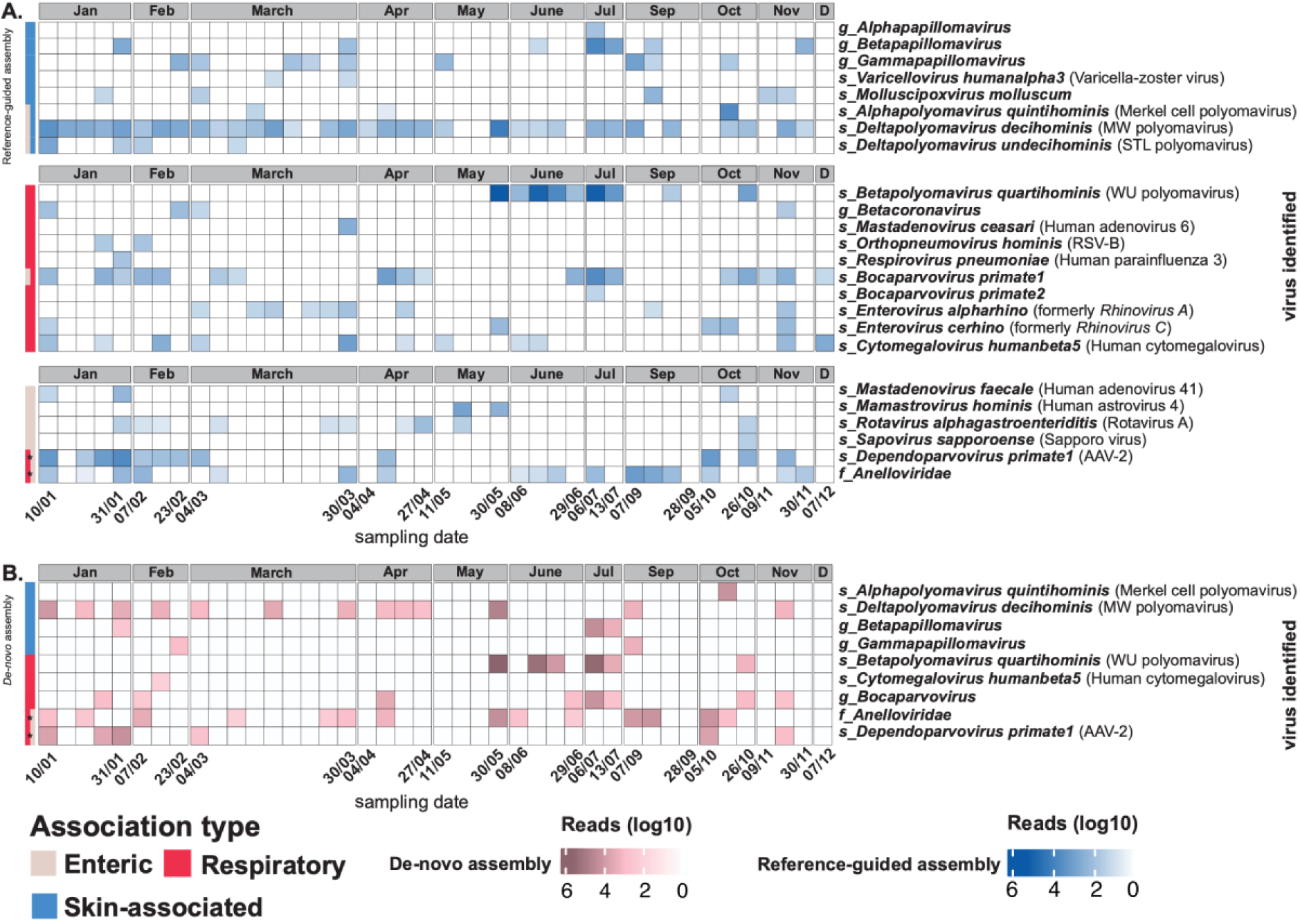
Human-associated viruses. On the x-axis, the first and last sample collection in each month is labelled. **A.** Human-associated viruses, identified by reference-based assembly. **B.** Human-associated viruses identified by de-novo assembly. Different taxonomical levels of identifications are abbreviated as follows: f: family g: genus, s: species. ***\*****AAV-2 is known to infect a wide range of human cells and anelloviruses do not have a clear host cell, however both are reported to shed from the enteric and respiratory tracts*.

Enteric virus reads detected in the air included rotavirus A (n=10), human astrovirus 4 (n=2), sapporo virus (n=1), and human adenovirus 41 (n=1). The highest horizontal genome coverage from strictly enteric pathogens was reached for Human astrovirus 4 with more than 70% coverage. While these pathogens are known to cause significant illness in young children, other viruses such as MW polyomavirus, STL polyomavirus, human bocavirus 1-3, anelloviruses, and Adeno-associated virus-2 (AAV-2) are described in stool without being associated with enteric diseases. AAV-2 was identified in 12 samples, and anelloviruses were detected in 21 samples. In four of these 21 samples, anelloviruses were only identified through *de novo* assembly, highlighting their high genetic variability, which may have complicated detection by reference-based approaches.

Notably, the most frequently identified virus was MW polyomavirus, present in 80.9% (34/42) of the samples throughout the year. This virus showed 98.8% nucleotide similarity to a strain reported to be shedding from the skin of healthy individuals in the USA^25^. Several skin-associated polyomaviruses were detected, including STL polyomavirus and Merkel Cell polyomavirus. Additionally, Varicella-Zoster Virus (VZV) and variety of human papillomaviruses (18 different types, including HPV-151, HPV-120, HPV-129, HPV-9, and HPV-76, detected in 14 samples) were identified.

### 3. Seasonality of respiratory viruses was reflected in the air

Examination of the seasonality of respiratory viruses with shotgun metagenomics revealed distinct patterns. Rhinoviruses were detected throughout the year, showing no clear seasonal pattern. Bocaparvoviruses were identified frequently in blocks of several consecutive positive samples (mid-January, April, end-June, mid-October), presumably representing outbreaks. In contrast, viruses with well-established seasonal patterns, such as coronaviruses and RSV, typically peak during the winter months^26,27^ and were only detected in the months of January, February, and November in our study (**Figure 1A**).

To increase the resolution for seasonality analyses, we used qPCR respiratory panel results from 90 samples (the same 42 samples as above, plus 48 additional indoor air samples from the same year and location*, see methods*). RSV was detected only from January to April (n=12, CT range: 32.0-39.1) and November to December (n=7, CT range: 32.9-35.3). Similarly, human parainfluenza virus 3 was found in January to March (n=12, CT range: 33.5-38.7) and once in May (n=1, CT: 35.3). Human coronaviruses (HKU-1 and OC43) were detected 42 times, mostly between January and March (n=31, CT range: 27.9-37.8) and in November to December (n=6, CT range: 27.9-37.8). Thus, both qPCR panels and shotgun metagenomics revealed distinct seasonality patterns for respiratory viruses in indoor air, particularly for RSV and human coronaviruses. Interestingly, Human parainfluenza virus 3 was detected in winter by indoor air sampling at daycare center, which is different than its defined peak in the literature, late spring and early summer. Furthermore, the respiratory qPCR panel and shotgun metagenomics did not detect any SARS-CoV-2, whereas the more sensitive TaqPath assay (*see methods*) detected SARS-CoV-2 in 45 out of 90 samples (**Supplementary information 2. S1**.).

### 4. Indoor air surveillance as a non-invasive way to follow temporal trends of rhinovirus and rotavirus A

A recent study revealed that human rhinoviruses cause up to two-thirds as many hospital admissions in infants as RSV^28^, while rotavirus remains the most common gastroenteritis cause in children younger than five-year-old worldwide. In our data RSV was only identified in two indoor samples using shotgun metagenomics, potentially because infants with more severe disease were kept away from the daycare center. On the other hand, rotavirus and rhinoviruses were consistently detected (**Figure 1.A**), and therefore, they were selected for a more thorough comparison to other openly available surveillance data .

For rhinovirus epidemiology, from patients with respiratory symptoms from the Leuven university hospital, which tested positive for enteroviruses (which includes rhinoviruses; *see methods*) using the respiratory qPCR panel, were used as a proxy (**Figure 2.A**, brown bars). The same qPCR panel was applied to all indoor air samples (n=90), revealing a 95.5% positivity rate (87/90) (indicated with “PCR #” **Figure 2.A**). Shotgun metagenomics identified rhinoviruses in 30.9% (13/42) of the samples and not every month, which is lower compared to qPCR panel tests. Average qPCR values per month showed similar trend to clinical cases between January-March, May-July and September-November. Despite low read counts (read counts can be found in **Supplementary Information 1**), our shotgun metagenomics data allowed identification and subtyping of rhinoviruses (**Figure 2.A**, heatmap).

**Figure 2.**
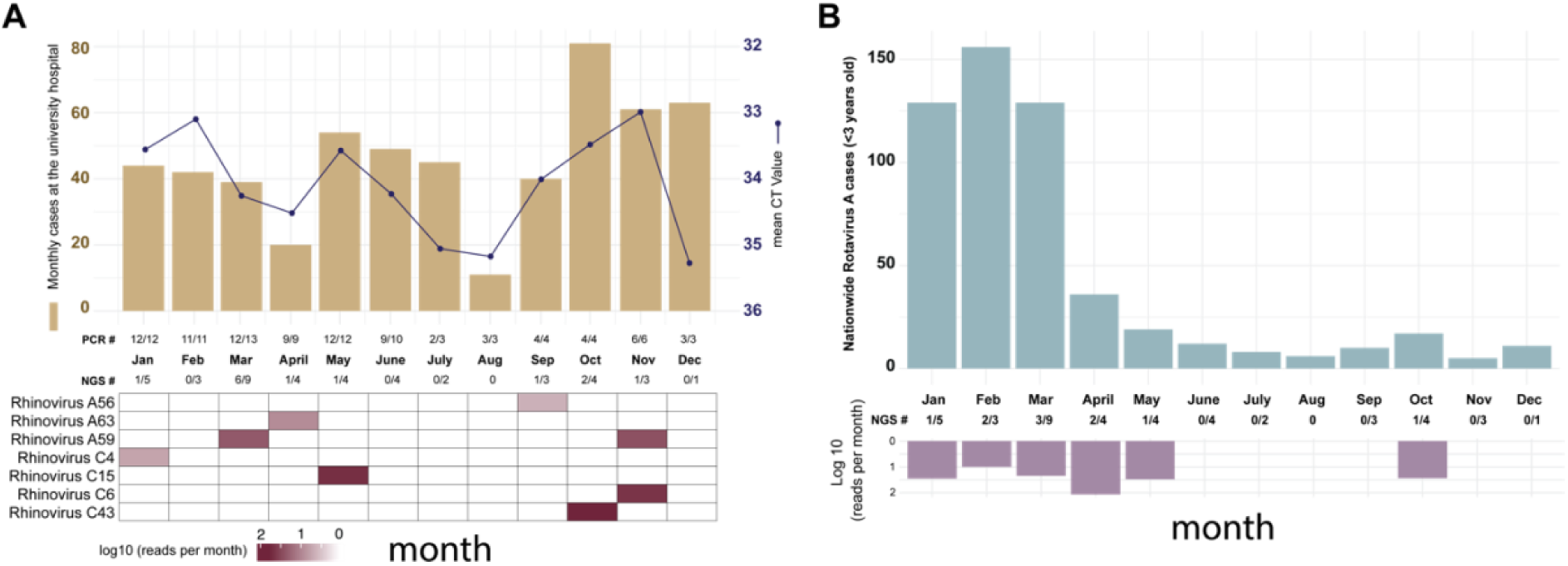
**A.** Monthly enterovirus positive (including rhinoviruses) cases at the university hospital laboratory (brown bars) and average qPCR CT values per month (black line) measured from indoor air samples collected at the daycare center coupled with heatmap (below) created using data from shotgun metagenomics. **B.** Monthly rotavirus cases nationwide coupled with inverted bar plot of aligned reads (log10 scale) per month.

To compare indoor air surveillance with actual rotavirus A circulation, nationwide data from the National Reference Center (NRC) for rotavirus A (*see methods*) among children younger than three *years old* were used (**Figure 2.B**). Unlike rhinoviruses, rotavirus A was not identified using the qPCR enteric panel (0/42), but it was detected in 10 samples (January to May and October) using untargeted shotgun metagenomics. These detections corresponded well with nationwide data and the seasonality reported by the NRC for rotaviruses in Belgium (**Figure 2.B**).

### 5. Untargeted shotgun metagenomics allows detailed phylogenetic analyses, and discrimination of distinct viral variants

In addition to the detection (absence or presence) of a particular pathogen, further molecular characterization of viral genomes can be important for public health purposes, such as subtyping or identifying mutations responsible for resistance against therapies. Therefore, we aimed to further leverage the obtained NGS data for the molecular characteristics of viral genomes found in indoor air samples. The data allowed us to reconstruct several partial and (near) complete genomes of several skin-associated (**Figure 3.A-C**) respiratory (**Figure 3.D-F**) and enteric viruses (**Figure 3. G-H**) as well as multiple near-complete genomes of AAV-2 (**Figure 3.I**). Moreover, various animal (bird and cat) viruses (**Figure 3.J-M**) and a complete genome of a novel densovirus were identified (see below). These obtained genomic data can further be used for phylogenetic analysis, as exemplified for the partial RSV genome data and the near-complete genomes of bocaviruses to reliable study their taxonomy and evolutionary relationships (**Figure 3.N-O**). The partial genome of RSV clustered closely with RSV-B sequences, whereas bocavirus genomes showed over 99% similarity to Human bocavirus 1 (OP255990) and human bocavirus 3 (NC_012564), reference sequences. Further examples of phylogenetic analyses for AAV-2, WU and MW polyomaviruses, HPV-151, and the partial genomes of felis domesticus papillomavirus, a novel densovirus, a divergent canary polyomavirus, and human astrovirus 4, are provided in **Supplementary information 2 S2**.

**Figure 3.**
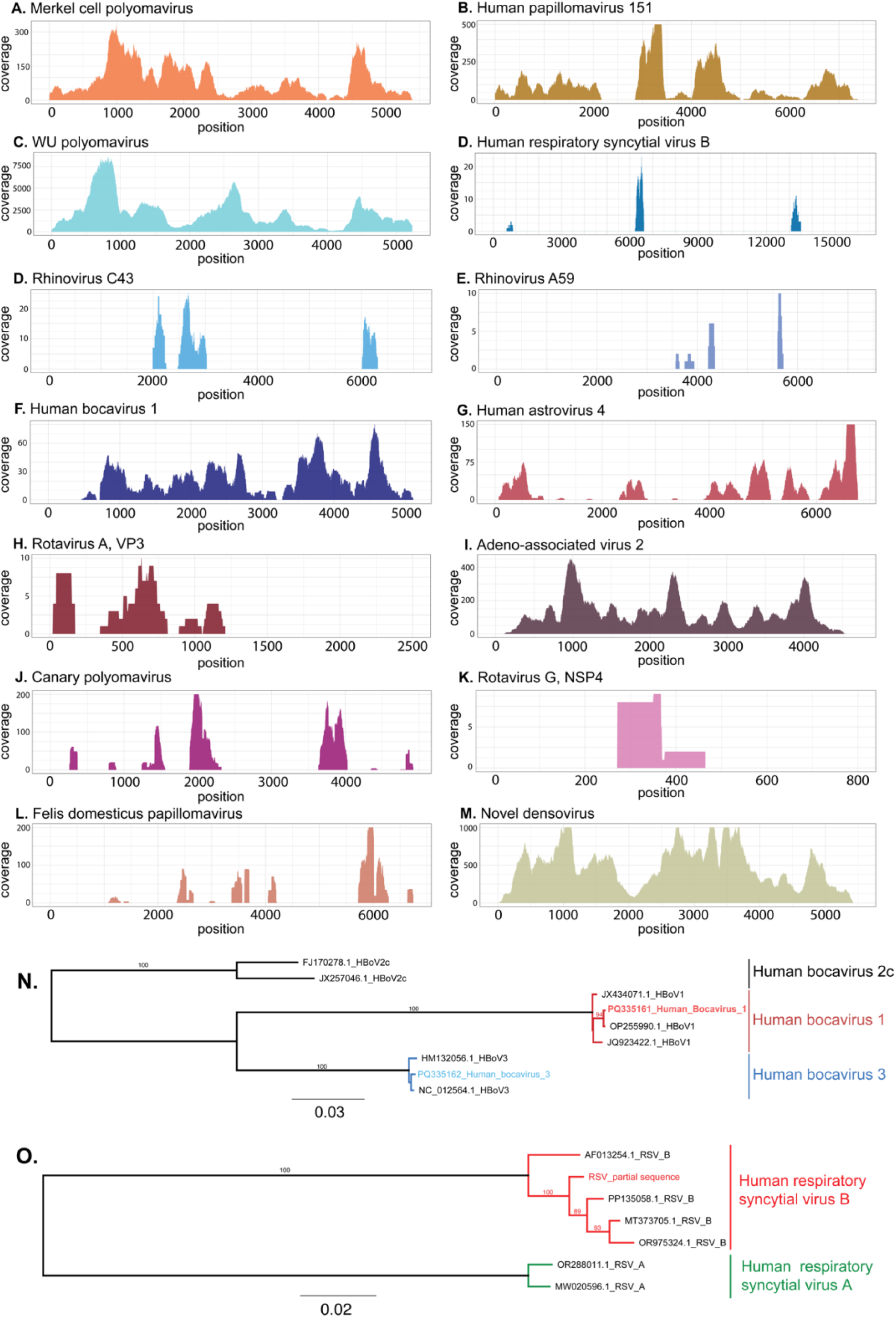
**A-M.** Coverage plots of viruses found in indoor air samples. The y-axis for each panel is different and shows the depth of coverage, while the x-axis shows nucleotide positions. **N-O.** Phylogenetic trees of 2 Human bocaviruses and RSV found in indoor air samples. Trees are constructed by the maximum likelihood method and numbers above the branch points denote the confidence level of the relationship of the paired sequences determined by bootstraps (n=100).

Interestingly, we detected WU polyomavirus in the daycare center for the first time at the end of May, and it was consistently present for nearly two months with high abundance, until the start of the summer break (**Figure 1.A**). This prompted us to investigate whether this prolonged detection was due to viral shedding of a single strain by one or more individuals or if there were multiple introductions of distinct viruses. Therefore, we reconstructed four near-complete genomes of WU polyomavirus from samples collected between the end of May and July (May 30th, June 15th, June 22^nd^ and July 6th). We show that the viral genomes collected at the first two time points are identical, whereas the third sample showed 12 mutations, suggesting the introduction of a second WU polyomavirus variant into the daycare center (mutations in different variants and read level analysis can be found in **Supplementary information 2. S3**). Finally, our in-depth read-level analysis suggests that both of these variants were present in the 4^th^ sample.

### 6. Untargeted shotgun metagenomics on indoor air samples can reflect temporal dynamics of both the indoor and outdoor environment

Next, we focused on the air virome linked to other entities such as plants, animals (including insects), and fungi. Although contigs identified as bacteriophages were also abundantly identified, due to their very short average contig length (∼1kb), we did not further explore them.

Untargeted deep sequencing, combined with reference-based assembly and *de novo* assembly (**Figure 5.A**), allowed us to detect insect and animal-associated viruses in 29 out of 42 samples (69%) (**Figure 4**). Avian species-infecting viruses, such as avian rotaviruses (detected in April, June, and October), and *Gammapolyomavirus Secanaria* (October) were detected. Additionally, honeybee-related viruses, such as Sacbrood virus and Moku virus (**Figure 4.A**), were identified in July and September, along with novel and divergent insect infecting densoviruses throughout the year (in 12 out of 42 samples). As expected, divergent (and novel) densoviruses were only identified by *de novo* assembly, while viruses with low read counts but high similarity to known references, such as avian rotaviruses, were only identified by reference-based assembly.

**Figure 4.**
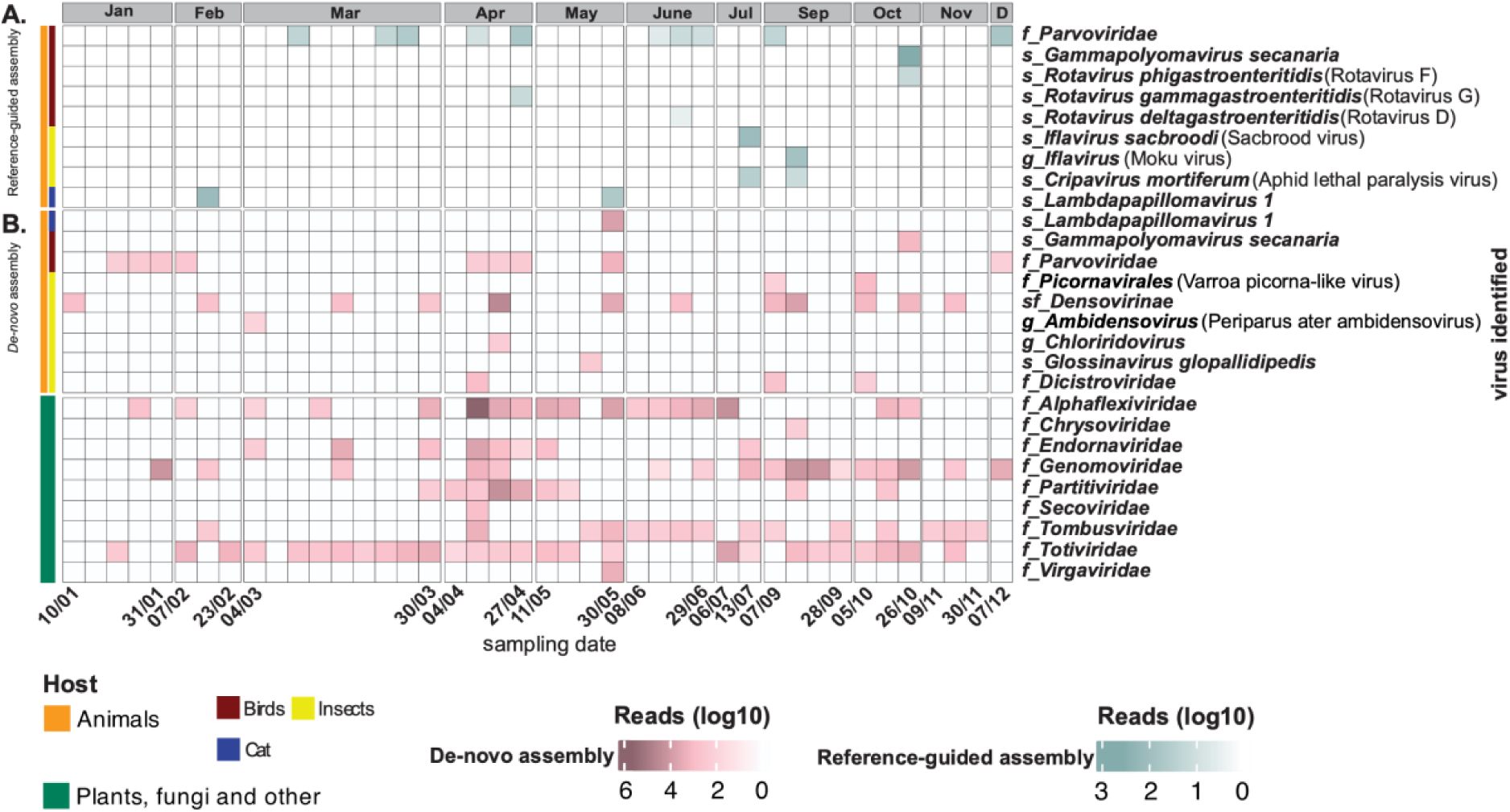
Animal (except humans) and plant viruses. On the x-axis, the first and last sample collection in each month is visible and the y-axis represents taxonomical level of viruses. **A.** Animal viruses identified by reference guided assembly. **B.** Animal and plant viruses identified by *de novo* assembly. Different taxonomical level of identifications abbreviated as follows: f: family g: genus, s: species, sf: subfamily.

**Figure 5.**
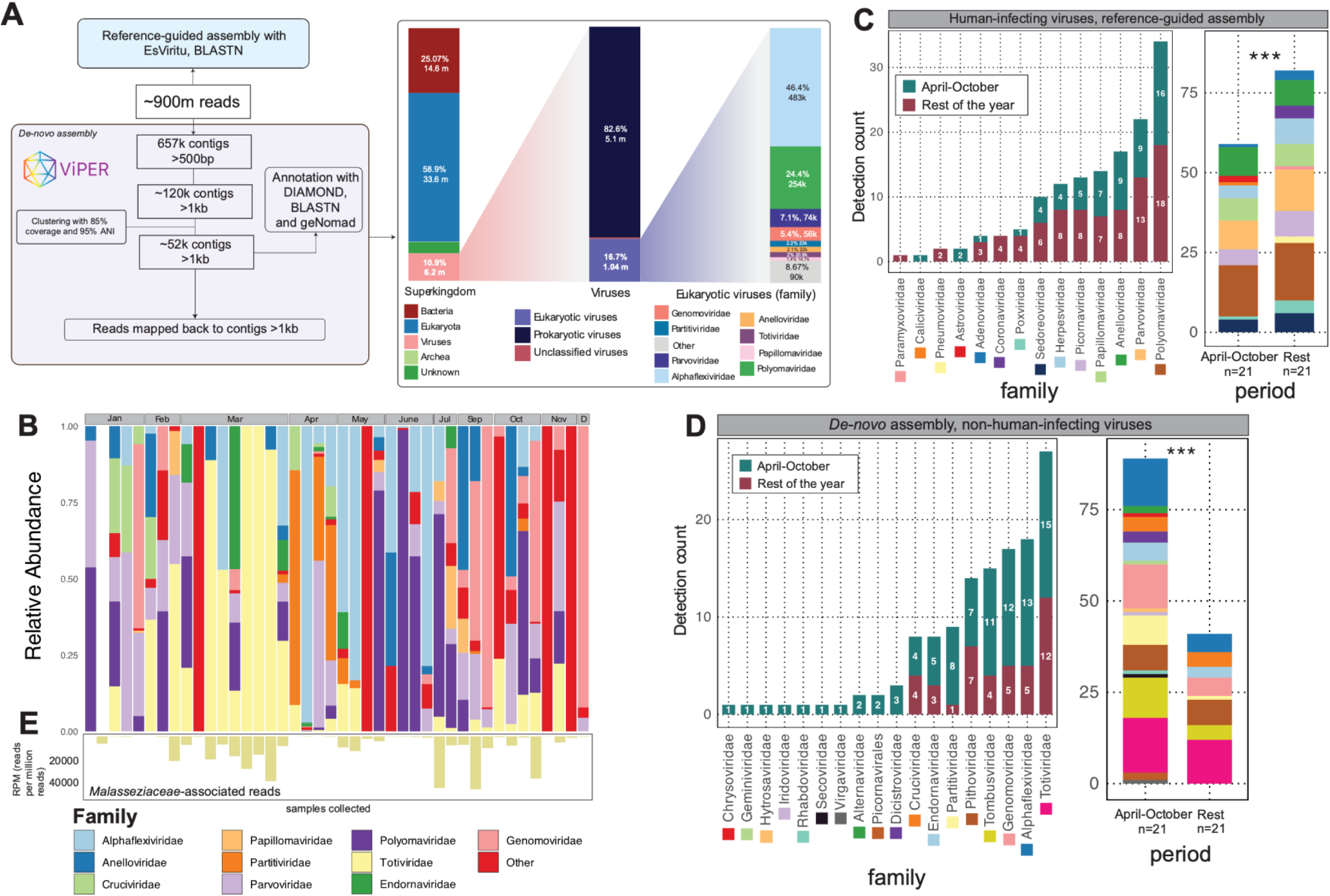
**A.** Bioinformatics pipeline used for reference-based and *de-novo* assembly-based virus identification. **B.** Relative abundance of different viral families detected in each sample. **C.** Number of detections of human-infecting viruses for each family (left panel) during period with open or closed windows. Aggregated detection counts of human-infecting viruses for the periods between April-October and rest of the year (right panel), X^2^=6.86, df=1, *p<0.01 (p= 0.0087).* **D.** Number of detections of non-human-infecting viruses for each family (left panel). Aggregated detection counts of human-infecting viruses for the periods between April-October and rest of the year (right panel), X^2^=33.985, df=1, *p<0.001.* **E.** Reads mapping back to contigs of family *Malasseziaceae*, as RPM (reads per million).

### 7. Non-human infecting viruses were more prevalent when the windows were open

A diverse array of plant and fungi infecting viruses were identified in 39/42 samples collected throughout the year. The identified viruses belonged to the families *Totiviridae* (predominantly fungi-infecting, n=27), *Tombusviridae* (plant-infecting, n=15), and 3 families of viruses known to infect both plants and fungi: *Genomoviridae* (n=17), *Partitiviridae* (n=9) and *Alphaflexiviridae* (n=18) (**Figure 4.B**, green panel and **Figure 5.B.**). The majority of animal-associated viruses were detected between April and October (**Figure 4.B**, orange panel), during the period when windows were open during the air collection periods (metadata can be found in **Supplementary information 2. S4**), suggesting a link to the outside environment. Next, we investigated the seasonality of human-infecting and non-human infecting viruses. Given that the daycare center typically kept its windows open from April to October, we categorized detections into two periods: April-October and the rest of the year. This categorization revealed a significantly higher prevalence of human-infecting viruses during the closed-window periods, while non-human-infecting viruses were more prevalent during the open-window period (*p*<0.01, **Figure 5.C-D**).

With respect to fungal viruses, we noted that some of the more abundantly identified viral genome sequences from the family *Totiviridae* were most closely related (∼80% amino acid identity) to viruses known to infect fungi associated with humans: “Geotrichum candidum infecting totivirus” and “Malassezia-restricta infecting totivirus” (**Figure 5.B**.). Overall, totiviruses were consistently detected in February-March (10 out of 12 samples) and July-October (7 out of 9 samples) (**Figure 5.B**.). Although our NGS protocol is aimed at enriching for virus sequences, the *de novo* approach also identified contigs classified as *Malasseziaecea*, and reads counts mapping to these contigs correlated strongly with the relative abundance of totiviruses (**Figure 5.B-E**).

## Discussion

Studies utilizing multi-pathogen qPCR and metagenomics on indoor air samples have highlighted the potential for tracking various viruses including respiratory, enteric and plant associated viruses^10–13,24,29^. Building on these studies, we employed a longitudinal approach over one year at a day care center using the NetoVIR protocol for VLP enrichment combined with short-read untargeted sequencing. Parents were advised to keep their children at home if they exhibited any symptoms of illness such as fever, runny nose, or diarrhea. This led to our study population consisting of “healthy” individuals. Throughout the 12-month period, we successfully detected a diverse array of human-associated viruses, including respiratory, enteric, and skin viruses, alongside non-human viruses linked to animals, plants, and fungi.

*Rosario et al.* identified Merkell cell polyomavirus, MW polyomavirus and papillomaviruses in air filter samples^29^. Similarly, polyomaviruses were the most abundant human viruses detected in our study. While WU polyomavirus found in our study has been reported in children with respiratory symptoms or immunocompromised individuals, other viruses have been reported among seemingly healthy individuals, such as Merkel cell polyomavirus and MW polyomavirus^30,31^. Although our understanding of polyomavirus transmission, cell tropism and its potential role in disease is currently limited, overall high prevalence and abundance of polyomaviruses suggests circulation of polyomaviruses among young individuals, as previously suggested^32^. Interestingly, alongside the human skin viruses, the most abundant fungal viruses were related to commensal human skin fungi from the family *Malasseziaceae*, providing evidence for the high abundance of human skin-associated microorganisms in the environment.

Recent studies have investigated the potential of indoor air surveillance to monitor specific viral pathogen dynamics and epidemiology in various settings, including hospitals, offices, and residential environments^10,12,24,33–36^. All these studies tracking epidemiology through air relied on targeted tests requiring specific viral genomic knowledge, while our study employed a non-targeted approach. The longitudinal design of our study, in combination with available epidemiological data from the Leuven university hospital, allowed us to use rhinovirus and rotavirus as examples to showcase the power of air sampling in epidemiological surveillance, especially for asymptomatic circulation in a setting with no clinically ill individuals. In addition, our NGS-based untargeted approach allowed us to subtype rhinoviruses, showing the co-circulation of various rhinoviruses throughout the year.

*Minor et al.* employed viral metagenomics for indoor air surveillance, detecting Influenza C in a preschool and providing genomic data for this frequently untested virus^24^. In our study, we have corroborated their findings by identifying multiple complete or partial genomes of viruses, such as Varicella zoster virus, papillomaviruses, and WU polyomavirus, which may be associated with respiratory or skin diseases but are not currently included in the routine qPCR panels for respiratory diseases in Belgian hospitals. Additionally, our study identified and was able to subtype bocavirus, rhinovirus, parainfluenza, and RSV-B, without requiring any additional molecular testing. In cases of high viral abundance (such as respiratory-associated WU polyomavirus), our methods are even able to distinguish the co-circulation of closely related variants. This underscores the potential of untargeted metagenomics in uncovering a broader range of viral diversity than targeted qPCR panels, highlighting its utility in monitoring both common and lesser-known viruses that may otherwise go undetected. Furthermore, utilizing *de novo* and reference-guided assembly in combination, we discovered novel and divergent viruses, demonstrating that we can use indoor air as a virus surveillance medium for pandemic preparedness.

Beyond surveillance of human viruses, indoor air and environmental surveillance with metagenomics could also be of interest for animal farms and greenhouses. *Kwok et al.* analyzed dust samples and pooled chicken feces from farms using shotgun metagenomics, identifying multiple viruses from the families *Parvoviridae, Picornaviridae, Caliciviridae,* and *Astroviridae* that matched those found in the feces from the same location^37^. In our study, we unexpectedly detected avian viruses (rotaviruses and a divergent polyomavirus), a cat papillomavirus, and several insect-infecting viruses, including a novel densovirus, in an animal-free environment. We propose that the animal viruses mentioned are highly resilient in the environment and may be transported indoors by insects, air currents, or even on human skin, where they can subsequently be shed. Additionally, the presence of multiple plant viruses demonstrates that indoor air can serve as an effective medium for detecting pathogens that typically require labor-intensive sampling from various crops. Therefore, we propose that indoor air surveillance in strategic animal and plant production locations can be invaluable for revealing epidemiological trends and potential outbreaks of viruses.

It was previously reported that indoor air shotgun metagenomics with other methods resulted in 1% to 3.2% of reads that could be assigned to viruses^38^, underscoring the low viral biomass of indoor air samples. However, using the NetoVIR protocol, 10.9% of reads mapped to viral contigs, primarily phages, aligning with earlier findings^39^. Interestingly, shotgun metagenomics identified a broader diversity of enteric viruses, such as rotavirus and sapovirus, than the enteric qPCR panel, whereas the respiratory qPCR panel detected rhinovirus, RSV, adenovirus, and parainfluenza more consistently than metagenomics. These differences likely reflect the distinct sensitivities of the qPCR panels, which are tailored for detecting specific viral loads in defined sample types. The viral load in indoor air is likely at or near detection thresholds, with commonly circulating human viruses comprising less than 0.005% of total reads, as reported by *Prussin et al*.^39^. In our study, we confirm this low viral load of human-associated viruses in indoor air with the exception of skin- and respiratory-associated polyomaviruses. Overall, we suggest that our method is robust for indoor air surveillance and combining various bioinformatics approaches can enhance our understanding of the air virome.

Given the longitudinal nature of our sample collection, it is possible to discuss seasonality of viruses. *Prussin II et al*. employed passive indoor air sampling and shotgun metagenomics throughout the year in a daycare center. They found human-associated viruses to be much more diverse and dominant in the winter, while the summertime virome contained a higher proportion and diversity of plant-associated viruses^39^. Similarly, we also observed a higher prevalence of animal- or plant-associated viruses when windows were open, particularly during the warmer months from April to October and respiratory and enteric viruses mostly during the wintertime when windows were closed. Therefore, we suggest that active indoor air sampling can reflect dynamics of both the inside and outside environment and can be a way to track indoor air quality.

Overall, untargeted viral metagenomics offers a strong and complementary tool for pathogen surveillance via indoor air, particularly in cases of suspected asymptomatic shedding. Future research should validate this approach at sentinel locations for skin, enteric, and respiratory viruses in targeted settings, ideally incorporating clinical infection data of individuals. Our findings underscore the ability of our method to investigate pathogenic animal and plant viruses, in relevant settings and natural environments, utilizing air sampling techniques and combining *de novo* and reference-based bioinformatics approaches.

Our study is subject to a number of limitations. Primarily, it was conducted at a single location, without access to information regarding the health status or medical history of individuals in the vicinity. Nonetheless, it was assumed that individuals were unlikely to exhibit severe respiratory or gastroenteritis-related symptoms, as it was recommended that families keep children at home if they displayed such symptoms. Secondly, our inability to acquire complete genomes for numerous viruses present, which may stem from methodological constraints of sequencing and low viral load in the air. Additionally, our results are not confirmed by culture-based techniques to determine if the detected viral genomes are indicative for infectious viruses. This would be important to know, with respect to the potential of airborne transmission of skin-associated and enteric viruses. On the other hand, our study has several significant strengths. Firstly, we employed a longitudinal approach over a 12-month period, allowing for monitoring of seasonal dynamics. Secondly, we utilized advanced viral enrichment methods, which enhanced our ability to detect even low-abundance viral species with greater sensitivity. Additionally, we conducted in-depth bioinformatics analysis, combining *de novo* assembly and reference-guided assembly techniques. This approach resulted in the identification of not only known human-associated viruses with very low abundance but also novel and divergent viruses, which can translate as “pathogen X” in the future, indicating its potential significance for pandemic preparedness efforts.

## Conclusion

Our study highlights indoor air sampling as a robust method for surveillance of both human-associated and non-human-associated viruses. We suggest that this method, implemented in sentinel locations and targeted populations, can be complementary to traditional patient-based surveillance methods.

## Methods

### Sample collection

Between January 2022 and December 2022, 90 indoor air samples were collected from a daycare center in Leuven, hosting children up to 3 years old (on average, 15 children and 3 adults). We aimed to select at least 4 samples from each month (weekly), however, after the respiratory PCR panel, sufficient material of 42 samples was available for further analysis with metagenomics and an enteric PCR panel. Sampling was done between 9.00-11.00 (AM) for 2 hours, which are active hours at the daycare center, with children being present in the main area. When the daycare center was closed, sample collection was not performed. For sampling, an AerosolSense active air sampler was used^10^. Ambient air was sampled with a rate of 200L/min through a vertical collection pipe and impacted onto the AerosolSense Capture Media (Thermo Fisher Scientific, Waltham, MA). CO_2_ concentrations were measured using a remote climate sensor (Elsys ERC CO2, Umeå, Sweden; placed next to the air sampler). Natural ventilation was also estimated by a Likert scale (no natural ventilation, one window open, door open, multiple windows open, door and window open).

### Sample processing

After removing the standard cartridges from the sampler, they were transported to the lab on the same day they were collected. Samples were dissolved in Universal Transport Medium (UTM), to be used for respiratory and enteric multiplex qPCR panels as well as metagenomics (see below). Samples were stored at 4 degrees until processing with the respiratory multiplex qPCR panel. If they required storage over the weekend, they were frozen at −80 degrees Celsius. Processing with the enteric multiplex qPCR panel and shotgun metagenomics was conducted in February 2023, on samples stored at -20 degrees Celsius, with 2 or less freeze thaw cycles. As negative controls, UTM (used to dissolve samples) was processed together and sequenced as deep as samples.

### Respiratory and enteric multiplex qPCR panels

Samples dissolved in UTM were subjected to multiplex quantitative PCR (qPCR) testing. The detailed protocols for these procedures have been previously described^10,40^. The respiratory panel is designed to detect twenty-two viruses and seven bacterial pathogens and was applied on 90 samples when they were collected. In contrast, the enteric panel detects seventeen non-viral pathogens (four parasites and thirteen bacteria) and six viruses, specifically Human adenovirus F40/F41, Norovirus GI, Norovirus GII, Rotavirus A, Astrovirus, and Sapovirus (genogroups GI, GII, GIV, and GV). The enteric panel was applied to the same 42 samples used for shotgun metagenomics (see below).

### Shotgun metagenomics

Samples were subjected to *Novel Enrichment Technique of Viromes* (NetoVIR)^41^. They were homogenized by vortexing for 10 seconds and homogenous samples were centrifuged at 17,000 g for 3 minutes and filtered with a 0.8µm PES (Polyethersulfone) centrifugal filter, in order to eliminate eukaryotic cells and bacteria. Free floating nucleic acids were removed by benzonase and micrococcal nuclease while being incubated for 2 hours at 37°C. DNA/RNA Amplification was done using a modified Whole Transcriptome Amplification 2 (WTA2) Kit to amplify both DNA and RNA. To obtain DNA amplification, the initial denaturation step was performed at 95°C instead of 70°C. Library preparation was done using a Nextera XT DNA Sample Preparation kit and sequencing was performed on an NovaSeq™ platform (Illumina).

### Clinical data collection

We collected the results of respiratory panel tests for enterovirus (including rhinovirus) performed at the clinical laboratory of University Hospitals Leuven during the same period as the air sample collection. The PCR target also included non-rhinovirus enteroviruses. However, we assumed that the majority of enteroviruses detected in individual respiratory samples were rhinoviruses. Clinical respiratory qRT-PCR panels are performed according to specific clinical indications. In immunocompetent individuals, these panels are utilized for respiratory infections requiring intensive care admission or those unresponsive to initial therapy for a respiratory infection. For immunocompromised patients, respiratory panels are more commonly used when lower respiratory infections are suspected.

For rotavirus A infections in the age group of 0 to 3 years old, we used data from the National Reference Center (NRC) for rotavirus in Belgium over the same period. Samples of the NRC were collected through nationwide laboratory system on a voluntary basis and under the regulations defined by the Royal Decree of 09/02/2011.

### Bioinformatics analyses

We employed *de novo* assembly and reference-guided assembly on the reads obtained from sequencing. Analyses were conducted on VSC KU Leuven (The Flemish Supercomputer Center) servers.

### Reference-guided assembly

To identify human and animal-associated viruses, we utilized EsViritu^42^ (v0.2.3) and BLASTN. EsViritu employs a database of all human and animal viruses in GenBank as of November 16th, 2022 (EsViritu DB v2.0.2), curated to include one representative genome per 95% average nucleotide identity cluster. Reads underwent quality control, adapter trimming, and mapping to reference sequences with 90% identity and 90% coverage. Viruses were considered present if the consensus sequence exceeded 200 base pairs or 10% of the reference genome and at least 2 reads mapped to it. Consensus sequences were obtained using Samtools consensus^43^ and subsequently verified using BLASTN. Reference-guided assembly only showed HPV-21 with 39 reads in one negative control, which resulted in a 236-nucleotide long consensus sequence, found to be exactly the same as in one other higher viral load sample with same virus, and was therefore classified as cross-contamination from this sample.

### *De-novo* assembly

For *de novo* assembly of viral genomes (or contigs), we utilized the Virome Paired-End Reads pipeline (ViPER)^44^. The pipeline processed quality-controlled and adapter-trimmed reads, performing *de novo* triple-assembly using metaSPAdes^45^. Contigs were then clustered at 95% identity and 85% coverage to reduce redundancy, first within individual samples and then among all samples. Reads were mapped back to contigs >1kbp, considering those with over 50% horizontal coverage present in each sample. Contig identification was subsequently performed using DIAMOND^46^ for eukaryotic viruses, geNomad^47^ for bacteriophages, and BLASTN for identification of genetic material associated to other entities (i.e., fungi, plant, human). For *de novo* assembly, all contigs present in negative controls were treated as contaminants and removed before downstream analyses.

### Statistical analyses

Statistical analyses for the difference of detections in open and closed window times of non-human infecting and human-infecting viruses were conducted using the chi-square test and performed in RStudio. The analysis assessed the association between seasonality and the presence of these viruses. A p-value of less than 0.05 was considered statistically significant.

### Data and code availability

Raw data, excluding human reads, is available at the Sequence Read Archive (SRA) with Bioproject number PRJNA1158979. Codes used to conduct the analyses and produce figures are available on https://github.com/Matthijnssenslab/Indoorair_daycare.

## Data Availability

Raw data, excluding human reads, is available at the Sequence Read Archive (SRA) with Bioproject number PRJNA1158979. Codes and processed data used to conduct the analyses and produce figures are available on https://github.com/Matthijnssenslab/Indoorair_daycare

## Acknowledgements

Mustafa Karatas is supported by a Research Foundation Flanders (FWO) fundamental research scholarship (number: 11P7I24N). We would like to thank Pieter Wets and Steven Traets for their help in collection of samples. Additionally, we thank the Rotavirus National Reference Center and National Reference Center for Respiratory Pathogens for their support throughout this research.

## Ethical approval

The study was approved by the Ethics Committee Research UZ/KU Leuven (S66518, B3222022000873). No informed consent was required from occupants of the sampled environments.

## Author contributions

Conceptualization was carried out by JM, EA, EK and MK. The project was managed by EK, LB, JR and EA. CE, BC, MK, and EK performed sample collection and processing. Data analysis, visualization, and drafting of the initial manuscript were conducted by MK. Supervision was provided by JM, EA, and EK. All authors reviewed and approved the final manuscript.

