## Supplementary information 1 for "Untargeted viral metagenomics of indoor air as a novel surveillance tool for respiratory, enteric and skin viruses"

| **Sample**  **Supplementary information 1.** Obtained consensus sequences and their information  **collection**  **date** | **accession** | **Covered**  **bases** | **Reference length** | **Completeness (%)** | **# reads aligned** | **Total # reads** | **family** | **species** | **subspecies** | **Respiratory panel CT-Value** | **Enteric panel CT-value** | **# people** | **Sample or Control** |
| --- | --- | --- | --- | --- | --- | --- | --- | --- | --- | --- | --- | --- | --- |
| **10/01/2022** | KY316160.1 | 385 | 34169 | 1.13 | 18 | 13662430 | Adenoviridae | Human mastadenovirus F | Human adenovirus 41 | 31.1 | 37,1 | 15 | Sample |
| **10/01/2022** | EF582385.1 | 245 | 7099 | 3.45 | 27 | 13662430 | Picornaviridae | Rhinovirus C | rhinovirus C4 | 32.2 | NA | 15 | Sample |
| **10/01/2022** | AB041007.1 | 915 | 3852 | 23.75 | 69 | 13662430 | Anelloviridae | Torque teno virus 1 | unclassified Torque teno virus 1 subspecies/strain | NA | NA | 15 | Sample |
| **10/01/2022** | DQ915164.2 | 534 | 31076 | 1.72 | 80 | 13662430 | Coronaviridae | Betacoronavirus 1 | HCoV- OC43 | 33.4 | NA | 15 | Sample |
| **10/01/2022** | KY490071.1 | 1270 | 235397 | 0.54 | 90 | 13662430 | Herpesviridae | Human betaherpesvirus 5 | Human betaherpesvirus 5 subspecies/strain | 31.1 | NA | 15 | Sample |
| **10/01/2022** | NC_007455.1 | 1150 | 5299 | 21.7 | 142 | 13662430 | Parvoviridae | Primate bocaparvovirus 1 | unclassified Primate bocaparvovirus 1 subspecies/strain | 29.3 | NA | 15 | Sample |
| **10/01/2022** | JX463183.1 | 3136 | 4776 | 65.66 | 619 | 13662430 | Polyomaviridae | Deltapolyomavirus undecihominis | STL polyomavirus | NA | NA | 15 | Sample |
| **10/01/2022** | AF043303.1 | 4180 | 4679 | 89.34 | 1583 | 13662430 | Parvoviridae | Adeno-associated dependoparvovirus A | adeno-associated virus 2 | NA | NA | 15 | Sample |
| **10/01/2022** | JX262162.1 | 4939 | 4939 | 100 | 2643 | 13662430 | Polyomaviridae | Deltapolyomavirus decihominis | unclassified Deltapolyomavirus decihominis subspecies/strain | NA | NA | 15 | Sample |
| **19/01/2022** | JQ898291.1 | 227 | 4927 | 4.61 | 16 | 15419422 | Polyomaviridae | Deltapolyomavirus decihominis | MW polyomavirus | NA | NA | 6 | Sample |
| **19/01/2022** | JX262162.1 | 2269 | 4939 | 45.94 | 461 | 15419422 | Polyomaviridae | Deltapolyomavirus decihominis | unclassified Deltapolyomavirus decihominis subspecies/strain | NA | NA | 6 | Sample |
| **24/01/2022** | AB054647.1 | 203 | 3790 | 5.36 | 2 | 6572238 | Anelloviridae | Torque teno virus 8 | unclassified Torque teno virus 8 subspecies/strain | NA | NA | 10 | Sample |
| **24/01/2022** | JQ898291.1 | 324 | 4927 | 6.58 | 3 | 6572238 | Polyomaviridae | Deltapolyomavirus decihominis | MW polyomavirus | NA | NA | 10 | Sample |
| **24/01/2022** | AF043303.1 | 773 | 4679 | 16.52 | 44 | 6572238 | Parvoviridae | Adeno-associated dependoparvovirus A | adeno-associated virus 2 | NA | NA | 10 | Sample |
| **24/01/2022** | JX262162.1 | 3972 | 4939 | 80.42 | 376 | 6572238 | Polyomaviridae | Deltapolyomavirus decihominis | unclassified Deltapolyomavirus decihominis subspecies/strain | NA | NA | 10 | Sample |
| **26/01/2022** | KY490071.1 | 231 | 235397 | 0.1 | 9 | 19014920 | Herpesviridae | Human betaherpesvirus 5 | unclassified Human betaherpesvirus 5 subspecies/strain | 29.6 | NA | 10 | Sample |
| **26/01/2022** | KY040275.1 | 1385 | 188253 | 0.74 | 17 | 19014920 | Poxviridae | Molluscum contagiosum virus | Molluscum contagiosum virus subtype 1 | NA | NA | 10 | Sample |
| **26/01/2022** | NC_001781.1 | 400 | 15225 | 2.63 | 46 | 19014920 | Pneumoviridae | Human orthopneumovirus | unclassified Human orthopneumovirus subspecies/strain | 35.7 | NA | 10 | Sample |
| **26/01/2022** | JX262162.1 | 1575 | 4939 | 31.89 | 260 | 19014920 | Polyomaviridae | Deltapolyomavirus decihominis | unclassified Deltapolyomavirus decihominis subspecies/strain | NA | NA | 10 | Sample |
| **26/01/2022** | NC_007455.1 | 2161 | 5299 | 40.78 | 292 | 19014920 | Parvoviridae | Primate bocaparvovirus 1 | unclassified Primate bocaparvovirus 1 subspecies/strain | 32.7 | NA | 10 | Sample |
| **26/01/2022** | AF043303.1 | 3073 | 4679 | 65.68 | 1229 | 19014920 | Parvoviridae | Adeno-associated dependoparvovirus A | adeno-associated virus 2 | NA | NA | 10 | Sample |
| **31/01/2022** | KC782520.2 | 163 | 2687 | 6.07 | 2 | 16984456 | Sedoreoviridae | Rotavirus A |  | NA | *Not detected* | 15 | Sample |
| **31/01/2022** | KC782515.2 | 119 | 1059 | 11.24 | 4 | 16984456 | Sedoreoviridae | Rotavirus A |  | NA | *Not detected* | 15 | Sample |
| **31/01/2022** | EF059923.1 | 372 | 2359 | 15.77 | 6 | 16984456 | Sedoreoviridae | Rotavirus A |  | NA | *Not detected* | 15 | Sample |
| **31/01/2022** | KC442988.1 | 192 | 2333 | 8.23 | 7 | 16984456 | Sedoreoviridae | Rotavirus A |  | NA | *Not detected* | 15 | Sample |
| **31/01/2022** | MG181725.1 | 286 | 2359 | 12.12 | 9 | 16984456 | Sedoreoviridae | Rotavirus A |  | NA | *Not detected* | 15 | Sample |
| **31/01/2022** | JX134046.1 | 356 | 2915 | 12.21 | 11 | 16984456 | Anelloviridae | TTV-like mini virus | unclassified TTV-like mini virus subspecies/strain | NA | NA | 15 | Sample |
| **31/01/2022** | AF261761.1 | 365 | 3736 | 9.77 | 36 | 16984456 | Anelloviridae | Torque teno virus 7 | unclassified Torque teno virus 7 subspecies/strain | NA | NA | 15 | Sample |
| **31/01/2022** | NC_007455.1 | 238 | 5299 | 4.49 | 38 | 16984456 | Parvoviridae | Primate bocaparvovirus 1 | unclassified Primate bocaparvovirus 1 subspecies/strain | 30.3 | NA | 15 | Sample |
| **31/01/2022** | JX463183.1 | 564 | 4776 | 11.81 | 41 | 16984456 | Polyomaviridae | Deltapolyomavirus undecihominis | STL polyomavirus | NA | NA | 15 | Sample |
| **31/01/2022** | EU326526.1 | 636 | 15462 | 4.11 | 70 | 16984456 | Paramyxoviridae | Human respirovirus 3 | unclassified Human respirovirus 3 subspecies/strain | 37.2 | NA | 15 | Sample |
| **31/01/2022** | AB060594.1 | 740 | 3234 | 22.88 | 130 | 16984456 | Anelloviridae | Torque teno virus 20 | unclassified Torque teno virus 20 subspecies/strain | NA | NA | 15 | Sample |
| **31/01/2022** | KY316160.1 | 2602 | 34169 | 7.62 | 184 | 16984456 | Adenoviridae | Human mastadenovirus F | Human adenovirus 41 | 29.1 | *Not detected* | 15 | Sample |
| **31/01/2022** | GQ845442.1 | 3512 | 7304 | 48.08 | 440 | 16984456 | Papillomaviridae | Betapapillomavirus 2 | Human papillomavirus 120 | NA | NA | 15 | Sample |
| **31/01/2022** | JX262162.1 | 4014 | 4939 | 81.27 | 850 | 16984456 | Polyomaviridae | Deltapolyomavirus decihominis | unclassified Deltapolyomavirus decihominis subspecies/strain | NA | NA | 15 | Sample |
| **31/01/2022** | AF043303.1 | 4436 | 4679 | 94.81 | 4396 | 16984456 | Parvoviridae | Adeno-associated dependoparvovirus A | adeno-associated virus 2 | NA | NA | 15 | Sample |
| **07/02/2022** | KJ753392.1 | 138 | 2530 | 5.45 | 2 | 9911346 | Sedoreoviridae | Rotavirus A |  | NA | *Not detected* | 15 | Sample |
| **07/02/2022** | KC782514.2 | 265 | 1566 | 16.92 | 4 | 9911346 | Sedoreoviridae | Rotavirus A |  | NA | *Not detected* | 15 | Sample |
| **07/02/2022** | JX262162.1 | 257 | 4939 | 5.2 | 5 | 9911346 | Polyomaviridae | Deltapolyomavirus decihominis | unclassified Deltapolyomavirus decihominis subspecies/strain | NA | NA | 15 | Sample |
| **07/02/2022** | JX463183.1 | 694 | 4776 | 14.53 | 36 | 9911346 | Polyomaviridae | Deltapolyomavirus undecihominis | STL polyomavirus | NA | NA | 15 | Sample |
| **07/02/2022** | AF043303.1 | 1127 | 4679 | 24.09 | 62 | 9911346 | Parvoviridae | Adeno-associated dependoparvovirus A | adeno-associated virus 2 | NA | NA | 15 | Sample |
| **07/02/2022** | NC_001781.1 | 1199 | 15225 | 7.88 | 65 | 9911346 | Pneumoviridae | Human orthopneumovirus | unclassified Human orthopneumovirus subspecies/strain | 32 | NA | 15 | Sample |
| **07/02/2022** | JQ898291.1 | 1263 | 4927 | 25.63 | 67 | 9911346 | Polyomaviridae | Deltapolyomavirus decihominis | MW polyomavirus | NA | NA | 15 | Sample |
| **07/02/2022** | NC_007455.1 | 2339 | 5299 | 44.14 | 205 | 9911346 | Parvoviridae | Primate bocaparvovirus 1 | unclassified Primate bocaparvovirus 1 subspecies/strain | 29.8 | NA | 15 | Sample |
| **07/02/2022** | MK212031.1 | 1433 | 2907 | 49.29 | 207 | 9911346 | Anelloviridae | TTV-like mini virus | unclassified TTV-like mini virus subspecies/strain | NA | NA | 15 | Sample |
| **21/02/2022** | JX040418.1 | 118 | 750 | 15.73 | 4 | 19682352 | Sedoreoviridae | Rotavirus A |  | NA | *Not detected* | 20 | Sample |
| **21/02/2022** | KU727766.1 | 563 | 5166 | 10.9 | 24 | 19682352 | Parvoviridae | Parus major densovirus | unclassified Parus major densovirus subspecies/strain | NA | NA | 20 | Sample |
| **21/02/2022** | NC_012564.1 | 391 | 5242 | 7.46 | 64 | 19682352 | Parvoviridae | Primate bocaparvovirus 1 | Human bocavirus 3 | 29.6 | NA | 20 | Sample |
| **21/02/2022** | AF043303.1 | 1394 | 4679 | 29.79 | 87 | 19682352 | Parvoviridae | Adeno-associated dependoparvovirus A | adeno-associated virus 2 | NA | NA | 20 | Sample |
| **21/02/2022** | EU796884.1 | 1333 | 7899 | 16.88 | 121 | 19682352 | Papillomaviridae | Dyothetapapillomavirus 1 | Felis domesticus papillomavirus 2 | NA | NA | 20 | Sample |
| **21/02/2022** | NC_007455.1 | 1293 | 5299 | 24.4 | 132 | 19682352 | Parvoviridae | Primate bocaparvovirus 1 | unclassified Primate bocaparvovirus 1 subspecies/strain | 29.6 | NA | 20 | Sample |
| **21/02/2022** | KY490071.1 | 5617 | 235397 | 2.39 | 345 | 19682352 | Herpesviridae | Human betaherpesvirus 5 | unclassified Human betaherpesvirus 5 subspecies/strain | 28.9 | NA | 20 | Sample |
| **21/02/2022** | JX262162.1 | 4059 | 4939 | 82.18 | 685 | 19682352 | Polyomaviridae | Deltapolyomavirus decihominis | unclassified Deltapolyomavirus decihominis subspecies/strain | NA | NA | 20 | Sample |
| **23/02/2022** | JQ898291.1 | 252 | 4927 | 5.11 | 10 | 13805648 | Polyomaviridae | Deltapolyomavirus decihominis | MW polyomavirus | NA | NA | 16 | Sample |
| **23/02/2022** | AF043303.1 | 857 | 4679 | 18.32 | 98 | 13805648 | Parvoviridae | Adeno-associated dependoparvovirus A | adeno-associated virus 2 | NA | NA | 16 | Sample |
| **23/02/2022** | KT779555.1 | 1102 | 29887 | 3.69 | 107 | 13805648 | Coronaviridae | Human coronavirus HKU1 | unclassified Human coronavirus HKU1 subspecies/strain | 27.9 | NA | 16 | Sample |
| **23/02/2022** | GU233853.1 | 1947 | 7219 | 26.97 | 306 | 13805648 | Papillomaviridae | Gammapapillomavirus 9 | Human papillomavirus type 129 | NA | NA | 16 | Sample |
| **23/02/2022** | JX262162.1 | 938 | 4939 | 18.99 | 483 | 13805648 | Polyomaviridae | Deltapolyomavirus decihominis | unclassified Deltapolyomavirus decihominis subspecies/strain | NA | NA | 16 | Sample |
| **04/03/2022** | JQ898291.1 | 391 | 4927 | 7.94 | 5 | 14696626 | Polyomaviridae | Deltapolyomavirus decihominis | MW polyomavirus | NA | NA | 17 | Sample |
| **04/03/2022** | KY629935.1 | 317 | 7130 | 4.45 | 15 | 14696626 | Picornaviridae | Rhinovirus A | rhinovirus A59 | 34.3 | NA | 17 | Sample |
| **04/03/2022** | KT779555.1 | 334 | 29887 | 1.12 | 19 | 14696626 | Coronaviridae | Human coronavirus HKU1 | unclassified Human coronavirus HKU1 subspecies/strain | 30.3 | NA | 17 | Sample |
| **04/03/2022** | KY490071.1 | 643 | 235397 | 0.27 | 26 | 14696626 | Herpesviridae | Human betaherpesvirus 5 | unclassified Human betaherpesvirus 5 subspecies/strain | 29.8 | NA | 17 | Sample |
| **04/03/2022** | KY040275.1 | 1039 | 188253 | 0.55 | 33 | 14696626 | Poxviridae | Molluscum contagiosum virus | Molluscum contagiosum virus subtype 1 | NA | NA | 17 | Sample |
| **04/03/2022** | GU233853.1 | 710 | 7219 | 9.84 | 59 | 14696626 | Papillomaviridae | Gammapapillomavirus 9 | Human papillomavirus type 129 | NA | NA | 17 | Sample |
| **04/03/2022** | AF043303.1 | 2569 | 4679 | 54.9 | 174 | 14696626 | Parvoviridae | Adeno-associated dependoparvovirus A | adeno-associated virus 2 | NA | NA | 17 | Sample |
| **04/03/2022** | JX262162.1 | 3188 | 4939 | 64.55 | 310 | 14696626 | Polyomaviridae | Deltapolyomavirus decihominis | unclassified Deltapolyomavirus decihominis subspecies/strain | NA | NA | 17 | Sample |
| **07/03/2022** | KM254174.1 | 236 | 3661 | 6.45 | 4 | 12801052 | Parvoviridae | Galliform chaphamaparvovirus 3 | chicken chapparvovirus HK | NA | NA | 23 | Sample |
| **07/03/2022** | AB054647.1 | 553 | 3790 | 14.59 | 8 | 12801052 | Anelloviridae | Torque teno virus 8 | unclassified Torque teno virus 8 subspecies/strain | NA | NA | 23 | Sample |
| **07/03/2022** | KC782516.2 | 117 | 1066 | 10.98 | 8 | 12801052 | Sedoreoviridae | Rotavirus A |  | NA | *Not detected* | 23 | Sample |
| **07/03/2022** | JX262162.1 | 439 | 4939 | 8.89 | 24 | 12801052 | Polyomaviridae | Deltapolyomavirus decihominis | unclassified Deltapolyomavirus decihominis subspecies/strain | NA | NA | 23 | Sample |
| **07/03/2022** | NC_012564.1 | 414 | 5242 | 7.9 | 43 | 12801052 | Parvoviridae | Primate bocaparvovirus 1 | Human bocavirus 3 | 34.4 | NA | 23 | Sample |
| **09/03/2022** | NC_012564.1 | 356 | 5242 | 6.79 | 14 | 16130390 | Parvoviridae | Primate bocaparvovirus 1 | Human bocavirus 3 | 33.6 | NA | 18 | Sample |
| **09/03/2022** | JX463183.1 | 713 | 4776 | 14.93 | 15 | 16130390 | Polyomaviridae | Deltapolyomavirus undecihominis | STL polyomavirus | NA | NA | 18 | Sample |
| **09/03/2022** | MG846442.1 | 496 | 4432 | 11.19 | 21 | 16130390 | Parvoviridae | Galliform chaphamaparvovirus 2 | Chicken chapparvovirus 2 | NA | NA | 18 | Sample |
| **09/03/2022** | JX262162.1 | 1072 | 4939 | 21.7 | 48 | 16130390 | Polyomaviridae | Deltapolyomavirus decihominis | unclassified Deltapolyomavirus decihominis subspecies/strain | NA | NA | 18 | Sample |
| **11/03/2022** | KY629935.1 | 557 | 7130 | 7.81 | 21 | 19895646 | Picornaviridae | Rhinovirus A | rhinovirus A59 | 31 | NA | 23 | Sample |
| **11/03/2022** | HM011556.1 | 838 | 5387 | 15.56 | 22 | 19895646 | Polyomaviridae | Alphapolyomavirus quintihominis | Merkel cell polyomavirus | NA | NA | 23 | Sample |
| **11/03/2022** | JX262162.1 | 1816 | 4939 | 36.77 | 99 | 19895646 | Polyomaviridae | Deltapolyomavirus decihominis | unclassified Deltapolyomavirus decihominis subspecies/strain | NA | NA | 23 | Sample |
| **14/03/2022** | MF898328.1 | 234 | 125095 | 0.19 | 10 | 12667232 | Herpesviridae | Human alphaherpesvirus 3 | unclassified Human alphaherpesvirus 3 subspecies/strain | NA | NA | 21 | Sample |
| **14/03/2022** | KY629935.1 | 420 | 7130 | 5.89 | 40 | 12667232 | Picornaviridae | Rhinovirus A | rhinovirus A59 | 33.1 | NA | 21 | Sample |
| **14/03/2022** | JX262162.1 | 4323 | 4939 | 87.53 | 1033 | 12667232 | Polyomaviridae | Deltapolyomavirus decihominis | unclassified Deltapolyomavirus decihominis subspecies/strain | NA | NA | 21 | Sample |
| **16/03/2022** | JX262162.1 | 461 | 4939 | 9.33 | 9 | 13284002 | Polyomaviridae | Deltapolyomavirus decihominis | unclassified Deltapolyomavirus decihominis subspecies/strain | NA | NA | 20 | Sample |
| **16/03/2022** | GU233853.1 | 2329 | 7219 | 32.26 | 76 | 13284002 | Papillomaviridae | Gammapapillomavirus 9 | Human papillomavirus type 129 | NA | NA | 20 | Sample |
| **21/03/2022** | KY629935.1 | 331 | 7130 | 4.64 | 12 | 19633702 | Picornaviridae | Rhinovirus A | rhinovirus A59 | 34.6 | NA | 22 | Sample |
| **21/03/2022** | MG846442.1 | 387 | 4432 | 8.73 | 18 | 19633702 | Parvoviridae | Galliform chaphamaparvovirus 2 | Chicken chapparvovirus 2 | NA | NA | 22 | Sample |
| **21/03/2022** | MF588730.1 | 212 | 7292 | 2.91 | 24 | 19633702 | Papillomaviridae | Gammapapillomavirus 18 | unclassified Gammapapillomavirus 18 subspecies/strain | NA | NA | 22 | Sample |
| **23/03/2022** | FJ169853.1 | 142 | 3305 | 4.3 | 8 | 16564138 | Sedoreoviridae | Rotavirus A |  | NA | *Not detected* | 18 | Sample |
| **23/03/2022** | KY629935.1 | 353 | 7130 | 4.95 | 18 | 16564138 | Picornaviridae | Rhinovirus A | rhinovirus A59 | 34.5 | NA | 18 | Sample |
| **23/03/2022** | KU569162.1 | 686 | 5154 | 13.31 | 38 | 16564138 | Parvoviridae | Galliform aveparvovirus 1 | unclassified Galliform aveparvovirus 1 subspecies/strain | NA | NA | 18 | Sample |
| **23/03/2022** | JX262162.1 | 1732 | 4939 | 35.07 | 108 | 16564138 | Polyomaviridae | Deltapolyomavirus decihominis | unclassified Deltapolyomavirus decihominis subspecies/strain | NA | NA | 18 | Sample |
| **30/03/2022** | KC782520.2 | 124 | 2687 | 4.61 | 6 | 19652094 | Sedoreoviridae | Rotavirus A |  |  | *Not detected* | 17 | Sample |
| **30/03/2022** | X74468.1 | 210 | 7412 | 2.83 | 13 | 19652094 | Papillomaviridae | Betapapillomavirus 2 | Human papillomavirus 15 | NA | NA | 17 | Sample |
| **30/03/2022** | MF898328.1 | 437 | 125095 | 0.35 | 14 | 19652094 | Herpesviridae | Human alphaherpesvirus 3 | unclassified Human alphaherpesvirus 3 subspecies/strain | NA | NA | 17 | Sample |
| **30/03/2022** | AB017613.1 | 246 | 3818 | 6.44 | 34 | 19652094 | Anelloviridae | Torque teno virus 16 | unclassified Torque teno virus 16 subspecies/strain | NA | NA | 17 | Sample |
| **30/03/2022** | GU233853.1 | 548 | 7219 | 7.59 | 58 | 19652094 | Papillomaviridae | Gammapapillomavirus 9 | Human papillomavirus type 129 | NA | NA | 17 | Sample |
| **30/03/2022** | FJ349096.1 | 2158 | 35758 | 6.04 | 71 | 19652094 | Adenoviridae | Human mastadenovirus C |  | 31.8 | NA | 17 | Sample |
| **30/03/2022** | KY629935.1 | 345 | 7130 | 4.84 | 85 | 19652094 | Picornaviridae | Rhinovirus A | rhinovirus A59 | *Not detected* | NA | 17 | Sample |
| **30/03/2022** | X74464.1 | 1700 | 7434 | 22.87 | 93 | 19652094 | Papillomaviridae | Betapapillomavirus 2 | Human papillomavirus 9 | NA | NA | 17 | Sample |
| **30/03/2022** | AC_000008.1 | 1784 | 35938 | 4.96 | 108 | 19652094 | Adenoviridae | Human mastadenovirus C |  | 31.8 | NA | 17 | Sample |
| **30/03/2022** | AF261761.1 | 870 | 3736 | 23.29 | 163 | 19652094 | Anelloviridae | Torque teno virus 7 | unclassified Torque teno virus 7 subspecies/strain | NA | NA | 17 | Sample |
| **30/03/2022** | AC_000017.1 | 2926 | 36001 | 8.13 | 175 | 19652094 | Adenoviridae | Human mastadenovirus C |  | 31.8 | NA | 17 | Sample |
| **30/03/2022** | JX262162.1 | 3172 | 4939 | 64.22 | 598 | 19652094 | Polyomaviridae | Deltapolyomavirus decihominis | unclassified Deltapolyomavirus decihominis subspecies/strain | NA | NA | 17 | Sample |
| **30/03/2022** | KY490071.1 | 12244 | 235397 | 5.2 | 998 | 19652094 | Herpesviridae | Human betaherpesvirus 5 | unclassified Human betaherpesvirus 5 subspecies/strain | 28.8 | NA | 17 | Sample |
| **04/04/2022** | JX262162.1 | 531 | 4939 | 10.75 | 17 | 15622914 | Polyomaviridae | Deltapolyomavirus decihominis | unclassified Deltapolyomavirus decihominis subspecies/strain | NA | NA | 15 | Sample |
| **08/04/2022** | HM011556.1 | 260 | 5387 | 4.83 | 3 | 12603994 | Polyomaviridae | Alphapolyomavirus quintihominis | Merkel cell polyomavirus | NA | NA | 16 | Sample |
| **08/04/2022** | MG846442.1 | 237 | 4432 | 5.35 | 7 | 12603994 | Parvoviridae | Galliform chaphamaparvovirus 2 | Chicken chapparvovirus 2 | NA | NA | 16 | Sample |
| **08/04/2022** | AB041962.1 | 263 | 2908 | 9.04 | 15 | 12603994 | Anelloviridae | Torque teno mini virus 5 | unclassified Torque teno mini virus 5 subspecies/strain | NA | NA | 16 | Sample |
| **08/04/2022** | AB017613.1 | 273 | 3818 | 7.15 | 20 | 12603994 | Anelloviridae | Torque teno virus 16 | unclassified Torque teno virus 16 subspecies/strain | NA | NA | 16 | Sample |
| **08/04/2022** | AF043303.1 | 1425 | 4679 | 30.46 | 133 | 12603994 | Parvoviridae | Adeno-associated dependoparvovirus A | adeno-associated virus 2 | NA | NA | 16 | Sample |
| **08/04/2022** | JX262162.1 | 2563 | 4939 | 51.89 | 334 | 12603994 | Polyomaviridae | Deltapolyomavirus decihominis | unclassified Deltapolyomavirus decihominis subspecies/strain | NA | NA | 16 | Sample |
| **08/04/2022** | NC_007455.1 | 4768 | 5299 | 89.98 | 1070 | 12603994 | Parvoviridae | Primate bocaparvovirus 1 | unclassified Primate bocaparvovirus 1 subspecies/strain | 26.9 | NA | 16 | Sample |
| **25/04/2022** | KU356638.1 | 190 | 751 | 25.3 | 5 | 9122068 | Sedoreoviridae | Rotavirus A |  | NA | *Not detected* | 22 | Sample |
| **25/04/2022** | KY490071.1 | 241 | 235397 | 0.1 | 8 | 9122068 | Herpesviridae | Human betaherpesvirus 5 | unclassified Human betaherpesvirus 5 subspecies/strain | 30 | NA | 22 | Sample |
| **25/04/2022** | KU727766.1 | 232 | 5166 | 4.49 | 10 | 9122068 | Parvoviridae | Parus major densovirus | unclassified Parus major densovirus subspecies/strain | NA | NA | 22 | Sample |
| **25/04/2022** | FJ445146.1 | 420 | 7141 | 5.88 | 30 | 9122068 | Picornaviridae | Rhinovirus A | rhinovirus A63 | 32.1 | NA | 22 | Sample |
| **25/04/2022** | NC_007455.1 | 844 | 5299 | 15.93 | 97 | 9122068 | Parvoviridae | Primate bocaparvovirus 1 | unclassified Primate bocaparvovirus 1 subspecies/strain | 28.5 | NA | 22 | Sample |
| **25/04/2022** | JX262162.1 | 3373 | 4939 | 68.29 | 334 | 9122068 | Polyomaviridae | Deltapolyomavirus decihominis | unclassified Deltapolyomavirus decihominis subspecies/strain | NA | NA | 22 | Sample |
| **27/04/2022** | KJ753392.1 | 190 | 2530 | 7.51 | 3 | 9993384 | Sedoreoviridae | Rotavirus A |  | NA | *Not detected* | 18 | Sample |
| **27/04/2022** | KC443588.1 | 155 | 2650 | 5.85 | 3 | 9993384 | Sedoreoviridae | Rotavirus A |  | NA | *Not detected* | 18 | Sample |
| **27/04/2022** | NC_021583.1 | 271 | 1295 | 20.93 | 4 | 9993384 | Sedoreoviridae | Rotavirus G |  | NA | NA | 18 | Sample |
| **27/04/2022** | KP882477.1 | 314 | 2508 | 12.52 | 4 | 9993384 | Sedoreoviridae | Rotavirus A |  | NA | *Not detected* | 18 | Sample |
| **27/04/2022** | KJ752082.1 | 132 | 834 | 15.83 | 8 | 9993384 | Sedoreoviridae | Rotavirus G |  | NA | NA | 18 | Sample |
| **27/04/2022** | KC178774.1 | 211 | 2687 | 7.85 | 10 | 9993384 | Sedoreoviridae | Rotavirus A |  | NA | *Not detected* | 18 | Sample |
| **27/04/2022** | NC_007455.1 | 294 | 5299 | 5.55 | 10 | 9993384 | Parvoviridae | Primate bocaparvovirus 1 | unclassified Primate bocaparvovirus 1 subspecies/strain | 29.9 | NA | 18 | Sample |
| **27/04/2022** | GU214704.1 | 241 | 5257 | 4.58 | 20 | 9993384 | Parvoviridae | Galliform aveparvovirus 1 | Chicken parvovirus ABU-P1 | NA | NA | 18 | Sample |
| **27/04/2022** | KC782520.2 | 485 | 2687 | 18.05 | 21 | 9993384 | Sedoreoviridae | Rotavirus A |  | NA | *Not detected* | 18 | Sample |
| **27/04/2022** | KY055429.1 | 925 | 2585 | 35.78 | 29 | 9993384 | Sedoreoviridae | Rotavirus A |  | NA | *Not detected* | 18 | Sample |
| **27/04/2022** | MG846442.1 | 347 | 4432 | 7.83 | 33 | 9993384 | Parvoviridae | Galliform chaphamaparvovirus 2 | Chicken chapparvovirus 2 | NA | NA | 18 | Sample |
| **27/04/2022** | KC782514.2 | 256 | 1566 | 16.35 | 40 | 9993384 | Sedoreoviridae | Rotavirus A |  | NA | *Not detected* | 18 | Sample |
| **27/04/2022** | JX262162.1 | 2467 | 4939 | 49.95 | 213 | 9993384 | Polyomaviridae | Deltapolyomavirus decihominis | unclassified Deltapolyomavirus decihominis subspecies/strain | NA | NA | 18 | Sample |
| **11/05/2022** | KY490071.1 | 201 | 235397 | 0.09 | 16 | 18147382 | Herpesviridae | Human betaherpesvirus 5 | unclassified Human betaherpesvirus 5 subspecies/strain | 29.8 | NA | 20 | Sample |
| **11/05/2022** | FJ755404.1 | 728 | 6771 | 10.75 | 34 | 18147382 | Astroviridae | Mamastrovirus 1 | Human astrovirus 1 Beijing/291/2007/CHN | NA | 37.9 | 20 | Sample |
| **11/05/2022** | JX262162.1 | 678 | 4939 | 13.73 | 38 | 18147382 | Polyomaviridae | Deltapolyomavirus decihominis | unclassified Deltapolyomavirus decihominis subspecies/strain | NA | NA | 20 | Sample |
| **11/05/2022** | GU233853.1 | 1166 | 7219 | 16.15 | 173 | 18147382 | Papillomaviridae | Gammapapillomavirus 9 | Human papillomavirus type 129 | NA | NA | 20 | Sample |
| **11/05/2022** | MF684776.1 | 1447 | 6803 | 21.27 | 247 | 18147382 | Astroviridae | Mamastrovirus 1 | Human astrovirus 5 | NA | 37.9 | 20 | Sample |
| **11/05/2022** | AY720891.1 | 2261 | 6723 | 33.63 | 493 | 18147382 | Astroviridae | Mamastrovirus 1 | Human astrovirus 4 | NA | 37.9 | 20 | Sample |
| **18/05/2022** | EF554148.1 | 143 | 3302 | 4.33 | 30 | 15571202 | Sedoreoviridae | Rotavirus A |  | NA | *Not detected* | 16 | Sample |
| **30/05/2022** | EU796884.1 | 296 | 7899 | 3.75 | 45 | 38552982 | Papillomaviridae | Dyothetapapillomavirus 1 | Felis domesticus papillomavirus 2 | NA | NA | 18 | Sample |
| **30/05/2022** | MF684776.1 | 472 | 6803 | 6.94 | 131 | 38552982 | Astroviridae | Mamastrovirus 1 | Human astrovirus 5 | NA | *Not detected* | 18 | Sample |
| **30/05/2022** | AY720891.1 | 637 | 6723 | 9.47 | 138 | 38552982 | Astroviridae | Mamastrovirus 1 | Human astrovirus 4 | NA | *Not detected* | 18 | Sample |
| **30/05/2022** | MG881840.1 | 599 | 6992 | 8.57 | 144 | 38552982 | Picornaviridae | Rhinovirus C | rhinovirus C15 | 32.3 | NA | 18 | Sample |
| **30/05/2022** | JX262162.1 | 4731 | 4939 | 95.79 | #### | 38552982 | Polyomaviridae | Deltapolyomavirus decihominis | unclassified Deltapolyomavirus decihominis subspecies/strain | NA | NA | 18 | Sample |
| **30/05/2022** | EF444549.1 | 5229 | 5229 | 100 | #### | 38552982 | Polyomaviridae | Betapolyomavirus quartihominis | WU Polyomavirus | NA | NA | 18 | Sample |
| **08/06/2022** | MK212031.1 | 344 | 2907 | 11.83 | 5 | 12284646 | Anelloviridae | TTV-like mini virus | unclassified TTV-like mini virus subspecies/strain | NA | NA | 18 | Sample |
| **08/06/2022** | KY490071.1 | 229 | 235397 | 0.1 | 9 | 12284646 | Herpesviridae | Human betaherpesvirus 5 | unclassified Human betaherpesvirus 5 subspecies/strain | 28.2 | NA | 18 | Sample |
| **08/06/2022** | JX262162.1 | 266 | 4939 | 5.39 | 16 | 12284646 | Polyomaviridae | Deltapolyomavirus decihominis | unclassified Deltapolyomavirus decihominis subspecies/strain | NA | NA | 18 | Sample |
| **08/06/2022** | KU727766.1 | 417 | 5166 | 8.07 | 40 | 12284646 | Parvoviridae | Parus major densovirus | unclassified Parus major densovirus subspecies/strain | NA | NA | 18 | Sample |
| **08/06/2022** | EF444549.1 | 1060 | 5229 | 20.27 | 84 | 12284646 | Polyomaviridae | Betapolyomavirus quartihominis | WU Polyomavirus | NA | NA | 18 | Sample |
| **15/06/2022** | MG846442.1 | 244 | 4432 | 5.51 | 3 | 17639852 | Parvoviridae | Galliform chaphamaparvovirus 2 | Chicken chapparvovirus 2 | NA | NA | 14 | Sample |
| **15/06/2022** | AX174942.1 | 205 | 3847 | 5.33 | 9 | 17639852 | Anelloviridae | Torque teno virus 22 | unclassified Torque teno virus 22 subspecies/strain | NA | NA | 14 | Sample |
| **15/06/2022** | KU727766.1 | 354 | 5166 | 6.85 | 13 | 17639852 | Parvoviridae | Parus major densovirus | unclassified Parus major densovirus subspecies/strain | NA | NA | 14 | Sample |
| **15/06/2022** | U31779.1 | 281 | 7779 | 3.61 | 14 | 17639852 | Papillomaviridae | Betapapillomavirus 1 | human papillomavirus 21 | NA | NA | 14 | Sample |
| **15/06/2022** | KY490071.1 | 312 | 235397 | 0.13 | 16 | 17639852 | Herpesviridae | Human betaherpesvirus 5 | unclassified Human betaherpesvirus 5 subspecies/strain | 29.9 | NA | 14 | Sample |
| **15/06/2022** | JX262162.1 | 324 | 4939 | 6.56 | 18 | 17639852 | Polyomaviridae | Deltapolyomavirus decihominis | unclassified Deltapolyomavirus decihominis subspecies/strain | NA | NA | 14 | Sample |
| **15/06/2022** | EF444549.1 | 5229 | 5229 | 100 | #### | 17639852 | Polyomaviridae | Betapolyomavirus quartihominis | WU Polyomavirus | NA | NA | 14 | Sample |
| **22/06/2022** | GU733444.1 | 131 | 2801 | 4.68 | 2 | 19761762 | Sedoreoviridae | Rotavirus D |  | NA | NA | 19 | Sample |
| **22/06/2022** | KU569162.1 | 441 | 5154 | 8.56 | 11 | 19761762 | Parvoviridae | Galliform aveparvovirus 1 | unclassified Galliform aveparvovirus 1 subspecies/strain | NA | NA | 19 | Sample |
| **22/06/2022** | KU727766.1 | 348 | 5166 | 6.74 | 35 | 19761762 | Parvoviridae | Parus major densovirus | unclassified Parus major densovirus subspecies/strain | NA | NA | 19 | Sample |
| **22/06/2022** | AB017613.1 | 670 | 3818 | 17.55 | 38 | 19761762 | Anelloviridae | Torque teno virus 16 | unclassified Torque teno virus 16 subspecies/strain | NA | NA | 19 | Sample |
| **22/06/2022** | JX262162.1 | 848 | 4939 | 17.17 | 41 | 19761762 | Polyomaviridae | Deltapolyomavirus decihominis | unclassified Deltapolyomavirus decihominis subspecies/strain | NA | NA | 19 | Sample |
| **22/06/2022** | EF444549.1 | 5229 | 5229 | 100 | 2653 | 19761762 | Polyomaviridae | Betapolyomavirus quartihominis | WU Polyomavirus | NA | NA | 19 | Sample |
| **29/06/2022** | KU727766.1 | 281 | 5166 | 5.44 | 4 | 12613618 | Parvoviridae | Parus major densovirus | unclassified Parus major densovirus subspecies/strain | NA | NA | 23 | Sample |
| **29/06/2022** | MG846442.1 | 394 | 4432 | 8.89 | 9 | 12613618 | Parvoviridae | Galliform chaphamaparvovirus 2 | Chicken chapparvovirus 2 | NA | NA | 23 | Sample |
| **29/06/2022** | EF444549.1 | 1186 | 5229 | 22.68 | 49 | 12613618 | Polyomaviridae | Betapolyomavirus quartihominis | WU Polyomavirus | NA | NA | 23 | Sample |
| **29/06/2022** | NC_007455.1 | 740 | 5299 | 13.96 | 56 | 12613618 | Parvoviridae | Primate bocaparvovirus 1 | unclassified Primate bocaparvovirus 1 subspecies/strain | 32.7 | NA | 23 | Sample |
| **29/06/2022** | NC_012564.1 | 2492 | 5242 | 47.54 | 227 | 12613618 | Parvoviridae | Primate bocaparvovirus 1 | Human bocavirus 3 | 32.7 | NA | 23 | Sample |
| **06/07/2022** | NC_012042.1 | 240 | 5196 | 4.62 | 22 | 23622894 | Parvoviridae | Primate bocaparvovirus 2 | Human bocavirus 2c PK | 30.6 | NA | 18 | Sample |
| **06/07/2022** | U37537.1 | 236 | 7868 | 3 | 75 | 23622894 | Papillomaviridae | Alphapapillomavirus 4 | Human papillomavirus type 57b | NA | NA | 18 | Sample |
| **06/07/2022** | AB017613.1 | 347 | 3818 | 9.09 | 122 | 23622894 | Anelloviridae | Torque teno virus 16 | unclassified Torque teno virus 16 subspecies/strain | NA | NA | 18 | Sample |
| **06/07/2022** | JX262162.1 | 1278 | 4939 | 25.88 | 266 | 23622894 | Polyomaviridae | Deltapolyomavirus decihominis | unclassified Deltapolyomavirus decihominis subspecies/strain | NA | NA | 18 | Sample |
| **06/07/2022** | NC_007455.1 | 2040 | 5299 | 38.5 | 739 | 23622894 | Parvoviridae | Primate bocaparvovirus 1 | unclassified Primate bocaparvovirus 1 subspecies/strain | 30.6 | NA | 18 | Sample |
| **06/07/2022** | NC_012564.1 | 3995 | 5242 | 76.21 | 3355 | 23622894 | Parvoviridae | Primate bocaparvovirus 1 | Human bocavirus 3 | 30.6 | NA | 18 | Sample |
| **06/07/2022** | FN677756.1 | 6648 | 7386 | 90.01 | 6002 | 23622894 | Papillomaviridae | Betapapillomavirus 2 | Human papillomavirus 151 | NA | NA | 18 | Sample |
| **06/07/2022** | EF444549.1 | 5229 | 5229 | 100 | #### | 23622894 | Polyomaviridae | Betapolyomavirus quartihominis | WU Polyomavirus | NA | NA | 18 | Sample |
| **13/07/2022** | KU727766.1 | 310 | 5166 | 6 | 12 | 23210736 | Parvoviridae | Parus major densovirus | unclassified Parus major densovirus subspecies/strain | NA | NA | 18 | Sample |
| **13/07/2022** | AF536531.1 | 360 | 9812 | 3.67 | 28 | 23210736 | Dicistroviridae | Aphid lethal paralysis virus | unclassified Aphid lethal paralysis virus subspecies/strain | NA | NA | 18 | Sample |
| **13/07/2022** | AF092924.1 | 606 | 8832 | 6.86 | 110 | 23210736 | Iflaviridae | Sacbrood virus | unclassified Sacbrood virus subspecies/strain | NA | NA | 18 | Sample |
| **13/07/2022** | JX262162.1 | 1205 | 4939 | 24.4 | 195 | 23210736 | Polyomaviridae | Deltapolyomavirus decihominis | unclassified Deltapolyomavirus decihominis subspecies/strain | NA | NA | 18 | Sample |
| **13/07/2022** | NC_007455.1 | 1780 | 5299 | 33.59 | 274 | 23210736 | Parvoviridae | Primate bocaparvovirus 1 | unclassified Primate bocaparvovirus 1 subspecies/strain | 30.3 | NA | 18 | Sample |
| **13/07/2022** | U31779.1 | 2190 | 7779 | 28.15 | 803 | 23210736 | Papillomaviridae | Betapapillomavirus 1 | human papillomavirus 21 | NA | NA | 18 | Sample |
| **13/07/2022** | EF444549.1 | 3756 | 5229 | 71.83 | 892 | 23210736 | Polyomaviridae | Betapolyomavirus quartihominis | WU Polyomavirus | NA | NA | 18 | Sample |
| **07/09/2022** | JQ898291.1 | 394 | 4927 | 8 | 6 | 27712580 | Polyomaviridae | Deltapolyomavirus decihominis | MW polyomavirus | NA | NA | 19 | Sample |
| **07/09/2022** | KU727766.1 | 355 | 5166 | 6.87 | 10 | 27712580 | Parvoviridae | Parus major densovirus | unclassified Parus major densovirus subspecies/strain | NA | NA | 19 | Sample |
| **07/09/2022** | AY629583.1 | 353 | 4682 | 7.54 | 20 | 27712580 | Parvoviridae | Avian dependoparvovirus 1 | Avian adeno-associated virus strain DA-1 | NA | NA | 19 | Sample |
| **07/09/2022** | AB041962.1 | 574 | 2908 | 19.74 | 26 | 27712580 | Anelloviridae | Torque teno mini virus 5 | unclassified Torque teno mini virus 5 subspecies/strain | NA | NA | 19 | Sample |
| **07/09/2022** | JX262162.1 | 2764 | 4939 | 55.96 | 611 | 27712580 | Polyomaviridae | Deltapolyomavirus decihominis | unclassified Deltapolyomavirus decihominis subspecies/strain | NA | NA | 19 | Sample |
| **07/09/2022** | MF588693.1 | 4516 | 7226 | 62.5 | 870 | 27712580 | Papillomaviridae | Gammapapillomavirus sp. | unclassified Gammapapillomavirus sp. subspecies/strain | NA | NA | 19 | Sample |
| **07/09/2022** | AB017613.1 | 2350 | 3818 | 61.55 | 1242 | 27712580 | Anelloviridae | Torque teno virus 16 | unclassified Torque teno virus 16 subspecies/strain | NA | NA | 19 | Sample |
| **14/09/2022** | FJ445140.1 | 291 | 7136 | 4.08 | 10 | 14298396 | Picornaviridae | Rhinovirus A | rhinovirus A56 | 32.7 | NA | 18 | Sample |
| **14/09/2022** | AF536531.1 | 213 | 9812 | 2.17 | 11 | 14298396 | Dicistroviridae | Aphid lethal paralysis virus | unclassified Aphid lethal paralysis virus subspecies/strain | NA | NA | 18 | Sample |
| **14/09/2022** | AB041962.1 | 210 | 2908 | 7.22 | 14 | 14298396 | Anelloviridae | Torque teno mini virus 5 | unclassified Torque teno mini virus 5 subspecies/strain | NA | NA | 18 | Sample |
| **14/09/2022** | KM085343.1 | 266 | 7278 | 3.65 | 20 | 14298396 | Papillomaviridae | Gammapapillomavirus 24 | Human papillomavirus 197 | NA | NA | 18 | Sample |
| **14/09/2022** | U31779.1 | 486 | 7779 | 6.25 | 51 | 14298396 | Papillomaviridae | Betapapillomavirus 1 | human papillomavirus 21 | NA | NA | 18 | Sample |
| **14/09/2022** | KU645789.1 | 595 | 10056 | 5.92 | 72 | 14298396 | Iflaviridae | Moku virus | unclassified Moku virus subspecies/strain | NA | NA | 18 | Sample |
| **14/09/2022** | AB060597.1 | 749 | 3246 | 23.07 | 204 | 14298396 | Anelloviridae | Torque teno virus 24 | unclassified Torque teno virus 24 subspecies/strain | NA | NA | 18 | Sample |
| **14/09/2022** | KY040275.1 | 1518 | 188253 | 0.81 | 212 | 14298396 | Poxviridae | Molluscum contagiosum virus | Molluscum contagiosum virus subtype 1 | NA | NA | 18 | Sample |
| **14/09/2022** | FR751463.1 | 1450 | 3725 | 38.93 | 332 | 14298396 | Anelloviridae | Torque teno virus | unclassified Torque teno virus subspecies/strain | NA | NA | 18 | Sample |
| **28/09/2022** | EF444549.1 | 329 | 5229 | 6.29 | 20 | 12518762 | Polyomaviridae | Betapolyomavirus quartihominis | WU Polyomavirus | NA | NA | 20 | Sample |
| **28/09/2022** | AB060597.1 | 711 | 3246 | 21.9 | 117 | 12518762 | Anelloviridae | Torque teno virus 24 | unclassified Torque teno virus 24 subspecies/strain | NA | NA | 20 | Sample |
| **28/09/2022** | JX262162.1 | 2346 | 4939 | 47.5 | 201 | 12518762 | Polyomaviridae | Deltapolyomavirus decihominis | unclassified Deltapolyomavirus decihominis subspecies/strain | NA | NA | 20 | Sample |
| **05/10/2022** | KU727766.1 | 394 | 5166 | 7.63 | 11 | 19405330 | Parvoviridae | Parus major densovirus | unclassified Parus major densovirus subspecies/strain | NA | NA | NA | Sample |
| **12/10/2022** | AB028668.1 | 281 | 3787 | 7.42 | 33 | 19253612 | Anelloviridae | Torque teno virus 15 | unclassified Torque teno virus 15 subspecies/strain | NA | NA | 20 | Sample |
| **12/10/2022** | KY369878.1 | 1096 | 7052 | 15.54 | 90 | 19253612 | Picornaviridae | Rhinovirus C | rhinovirus C43 | 30.3 | NA | 20 | Sample |
| **12/10/2022** | AF043303.1 | 3762 | 4679 | 80.4 | 1444 | 19253612 | Parvoviridae | Adeno-associated dependoparvovirus A | adeno-associated virus 2 | NA | NA | 20 | Sample |
| **19/10/2022** | KY316160.1 | 393 | 34169 | 1.15 | 27 | 20826006 | Adenoviridae | Human mastadenovirus F | Human adenovirus 41 | 32.8 | 37.2 | NA | Sample |
| **19/10/2022** | NC_007455.1 | 548 | 5299 | 10.34 | 44 | 20826006 | Parvoviridae | Primate bocaparvovirus 1 | unclassified Primate bocaparvovirus 1 subspecies/strain | 31.2 | NA | NA | Sample |
| **19/10/2022** | MF588728.1 | 994 | 7209 | 13.79 | 46 | 20826006 | Papillomaviridae | Gammapapillomavirus 16 | unclassified Gammapapillomavirus 16 subspecies/strain | NA | NA | NA | Sample |
| **19/10/2022** | KU727766.1 | 451 | 5166 | 8.73 | 46 | 20826006 | Parvoviridae | Parus major densovirus | unclassified Parus major densovirus subspecies/strain | NA | NA | NA | Sample |
| **19/10/2022** | JX262162.1 | 758 | 4939 | 15.35 | 58 | 20826006 | Polyomaviridae | Deltapolyomavirus decihominis | unclassified Deltapolyomavirus decihominis subspecies/strain | NA | NA | NA | Sample |
| **19/10/2022** | KY369878.1 | 1092 | 7052 | 15.48 | 143 | 20826006 | Picornaviridae | Rhinovirus C | rhinovirus C43 | 32.1 | NA | NA | Sample |
| **19/10/2022** | HM011556.1 | 5379 | 5387 | 99.85 | 4107 | 20826006 | Polyomaviridae | Alphapolyomavirus quintihominis | Merkel cell polyomavirus | NA | NA | NA | Sample |
| **26/10/2022** | FN665693.1 | 113 | 1356 | 8.33 | 13 | 21012810 | Sedoreoviridae | Rotavirus A |  | NA | *Not detected* | 23 | Sample |
| **26/10/2022** | KU356605.1 | 162 | 751 | 21.57 | 14 | 21012810 | Sedoreoviridae | Rotavirus A |  | NA | *Not detected* | 23 | Sample |
| **26/10/2022** | JQ919995.1 | 152 | 2769 | 5.49 | 14 | 21012810 | Sedoreoviridae | Rotavirus F |  | NA | NA | 23 | Sample |
| **26/10/2022** | X86560.1 | 212 | 7431 | 2.85 | 14 | 21012810 | Caliciviridae | Sapporo virus | Sapporo virus-Manchester | NA | Not detected | 23 | Sample |
| **26/10/2022** | KP298674.1 | 400 | 7429 | 5.38 | 16 | 21012810 | Caliciviridae | Sapporo virus | Sapovirus Hu/GI.1/Seoul/ROK62/2013/KOR | NA | Not detected | 23 | Sample |
| **26/10/2022** | JX262162.1 | 796 | 4939 | 16.12 | 102 | 21012810 | Polyomaviridae | Deltapolyomavirus decihominis | unclassified Deltapolyomavirus decihominis subspecies/strain | NA | NA | 23 | Sample |
| **26/10/2022** | AF043303.1 | 1598 | 4679 | 34.15 | 280 | 21012810 | Parvoviridae | Adeno-associated dependoparvovirus A | adeno-associated virus 2 | NA | NA | 23 | Sample |
| **26/10/2022** | GU345044.1 | 952 | 5421 | 17.56 | 361 | 21012810 | Polyomaviridae | Gammapolyomavirus secanaria | Canary polyomavirus | NA | NA | 23 | Sample |
| **26/10/2022** | NC_007455.1 | 1948 | 5299 | 36.76 | 394 | 21012810 | Parvoviridae | Primate bocaparvovirus 1 | unclassified Primate bocaparvovirus 1 subspecies/strain | 29.5 | NA | 23 | Sample |
| **26/10/2022** | EF444549.1 | 3494 | 5229 | 66.82 | 478 | 21012810 | Polyomaviridae | Betapolyomavirus quartihominis | WU Polyomavirus | NA | NA | 23 | Sample |
| **09/11/2022** | NC_007455.1 | 403 | 5299 | 7.61 | 22 | 13856312 | Parvoviridae | Primate bocaparvovirus 1 | unclassified Primate bocaparvovirus 1 subspecies/strain | 30.2 | NA | NA | Sample |
| **09/11/2022** | KU727766.1 | 451 | 5166 | 8.73 | 36 | 13856312 | Parvoviridae | Parus major densovirus | unclassified Parus major densovirus subspecies/strain | NA | NA | NA | Sample |
| **09/11/2022** | KY040275.1 | 1091 | 188253 | 0.58 | 40 | 13856312 | Poxviridae | Molluscum contagiosum virus | Molluscum contagiosum virus subtype 1 | NA | NA | NA | Sample |
| **16/11/2022** | JX134046.1 | 218 | 2915 | 7.48 | 12 | 22190290 | Anelloviridae | TTV-like mini virus | unclassified TTV-like mini virus subspecies/strain | NA | NA | NA | Sample |
| **16/11/2022** | KU727766.1 | 368 | 5166 | 7.12 | 19 | 22190290 | Parvoviridae | Parus major densovirus | unclassified Parus major densovirus subspecies/strain | NA | NA | NA | Sample |
| **16/11/2022** | KY040275.1 | 1059 | 188253 | 0.56 | 24 | 22190290 | Poxviridae | Molluscum contagiosum virus | Molluscum contagiosum virus subtype 1 | NA | NA | NA | Sample |
| **16/11/2022** | KT779555.1 | 492 | 29887 | 1.65 | 41 | 22190290 | Coronaviridae | Human coronavirus HKU1 | unclassified Human coronavirus HKU1 subspecies/strain | NA | NA | NA | Sample |
| **16/11/2022** | KY629935.1 | 335 | 7130 | 4.7 | 75 | 22190290 | Picornaviridae | Rhinovirus A | rhinovirus A59 | 32.9 | NA | NA | Sample |
| **16/11/2022** | EF582387.1 | 1059 | 7086 | 14.94 | 129 | 22190290 | Picornaviridae | Rhinovirus C | rhinovirus C6 | 32.9 | NA | NA | Sample |
| **16/11/2022** | KY490071.1 | 1822 | 235397 | 0.77 | 147 | 22190290 | Herpesviridae | Human betaherpesvirus 5 | unclassified Human betaherpesvirus 5 subspecies/strain | 32.8 | NA | NA | Sample |
| **16/11/2022** | AF043303.1 | 2228 | 4679 | 47.62 | 257 | 22190290 | Parvoviridae | Adeno-associated dependoparvovirus A | adeno-associated virus 2 | NA | NA | NA | Sample |
| **16/11/2022** | NC_007455.1 | 2393 | 5299 | 45.16 | 322 | 22190290 | Parvoviridae | Primate bocaparvovirus 1 | unclassified Primate bocaparvovirus 1 subspecies/strain | 32.4 | NA | NA | Sample |
| **16/11/2022** | JX262162.1 | 3207 | 4939 | 64.93 | 481 | 22190290 | Polyomaviridae | Deltapolyomavirus decihominis | unclassified Deltapolyomavirus decihominis subspecies/strain | NA | NA | NA | Sample |
| **23/11/2022** | AB041007.1 | 322 | 3852 | 8.36 | 13 | 14766256 | Anelloviridae | Torque teno virus 1 | unclassified Torque teno virus 1 subspecies/strain | NA | NA | NA | Sample |
| **23/11/2022** | AB038621.1 | 413 | 3676 | 11.24 | 16 | 14766256 | Anelloviridae | Torque teno virus 29 | unclassified Torque teno virus 29 subspecies/strain | NA | NA | NA | Sample |
| **23/11/2022** | JX262162.1 | 416 | 4939 | 8.42 | 18 | 14766256 | Polyomaviridae | Deltapolyomavirus decihominis | unclassified Deltapolyomavirus decihominis subspecies/strain | NA | NA | NA | Sample |
| **23/11/2022** | AX025718.1 | 370 | 3313 | 11.17 | 19 | 14766256 | Anelloviridae | Torque teno virus 18 | unclassified Torque teno virus 18 subspecies/strain | NA | NA | NA | Sample |
| **23/11/2022** | KU727766.1 | 467 | 5166 | 9.04 | 28 | 14766256 | Parvoviridae | Parus major densovirus | unclassified Parus major densovirus subspecies/strain | NA | NA | NA | Sample |
| **23/11/2022** | Y15174.1 | 1846 | 7549 | 24.45 | 264 | 14766256 | Papillomaviridae | Betapapillomavirus 3 | human papillomavirus 76 | NA | NA | NA | Sample |
| **07/12/2022** | NC_007455.1 | 610 | 5299 | 11.51 | 15 | 11691674 | Parvoviridae | Primate bocaparvovirus 1 | unclassified Primate bocaparvovirus 1 subspecies/strain | 30 | NA | NA | Sample |
| **07/12/2022** | MG846442.1 | 280 | 4432 | 6.32 | 57 | 11691674 | Parvoviridae | Galliform chaphamaparvovirus 2 | Chicken chapparvovirus 2 | NA | NA | NA | Sample |
| **07/12/2022** | KY490071.1 | 3064 | 235397 | 1.3 | 403 | 11691674 | Herpesviridae | Human betaherpesvirus 5 | unclassified Human betaherpesvirus 5 subspecies/strain | 32.2 | NA | NA | Sample |
| **NA** | U31779.1 | 233 | 7779 | 3 | 39 | 14703762 | Papillomaviridae | Betapapillomavirus 1 | human papillomavirus 21 | NA | NA | NA | Control |
