## Supplementary information 2 for "Untargeted viral metagenomics of indoor air as a novel surveillance tool for respiratory, enteric and skin viruses"

**Supplementary information 2. S1.** Monthly average PCR CT values per pathogen. Pathogens detected less than 10 times were excluded, blue shade shows confidence interval and was only calculated when more than 6 samples were positive for that month.

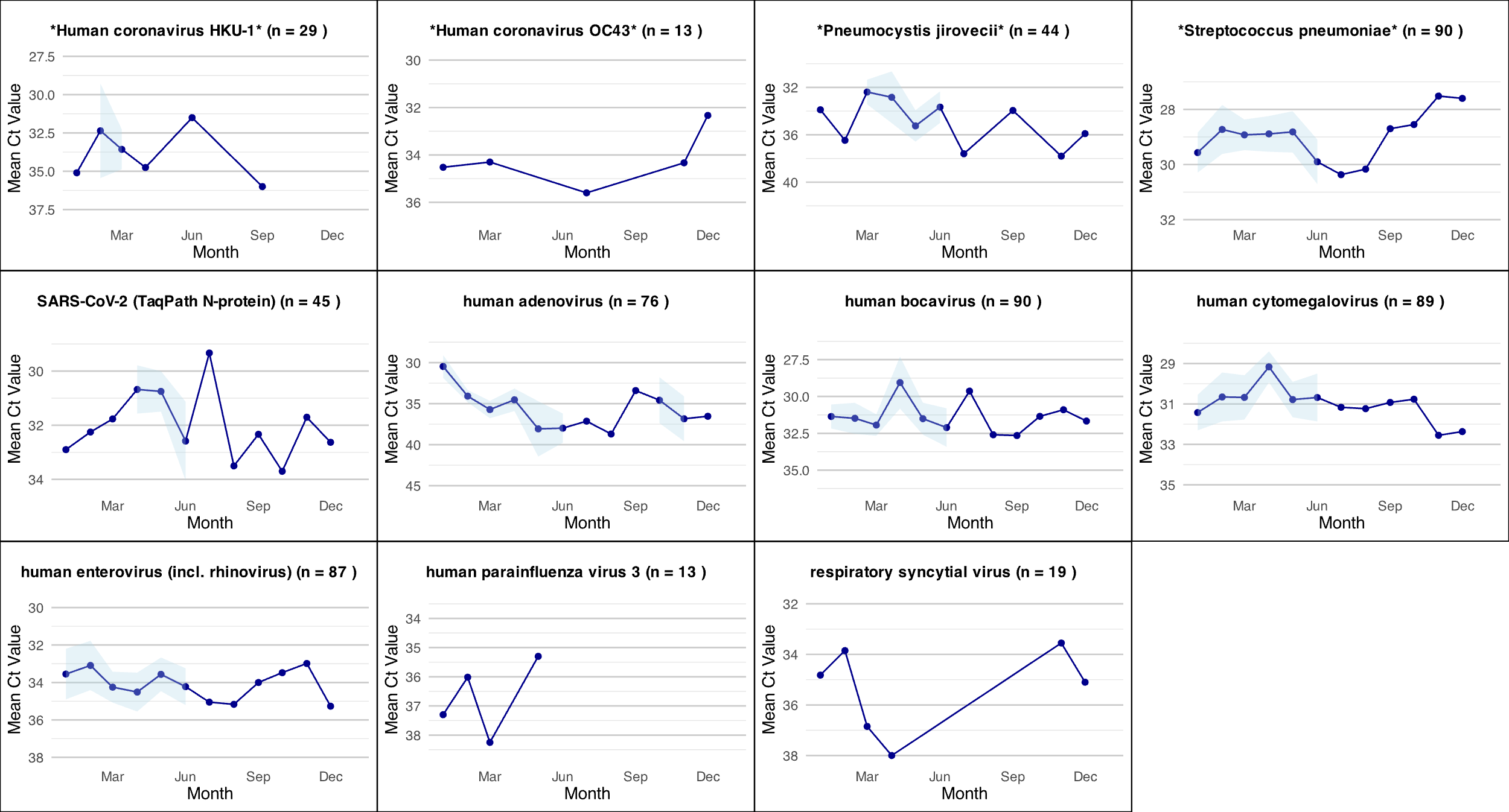

**Supplementary information 2.S2.** Phylogenetic trees of a divergent canary polyomavirus, felis domesticus papillomavirus, Adeno-associated virus 2, human astrovirus, MW polyomavirus, WU polyomavirus and novel densovirus.

*Canary polyomavirus*

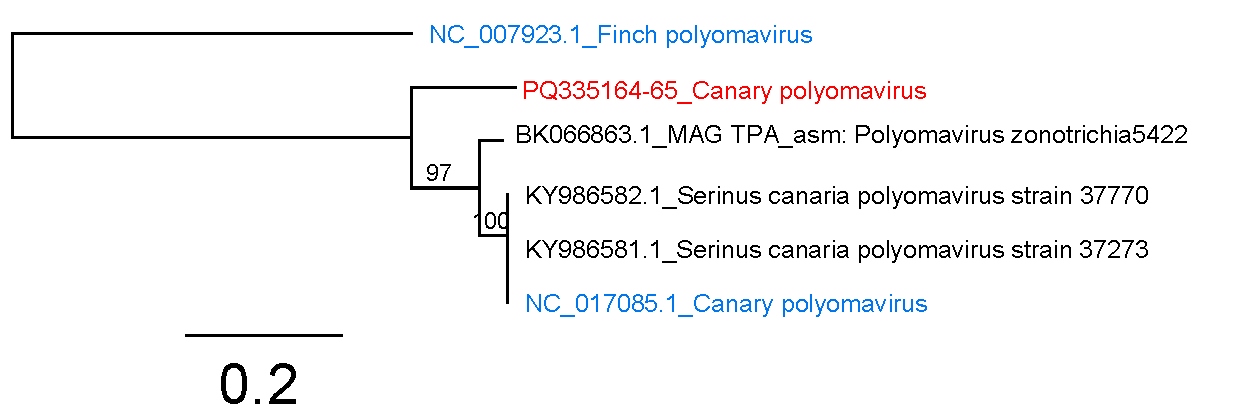

*Felis domesticus papillomavirus*

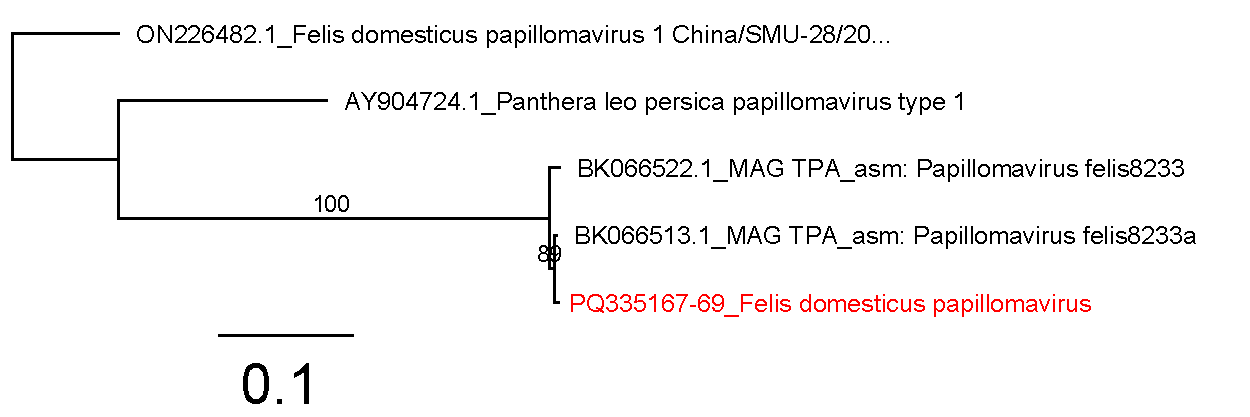

*Adeno-associated virus 2*

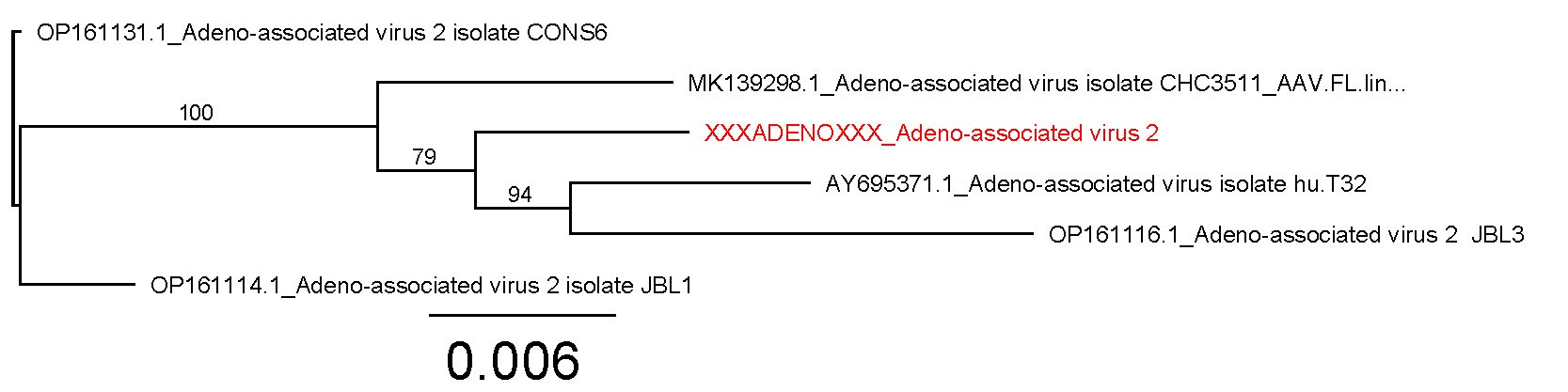

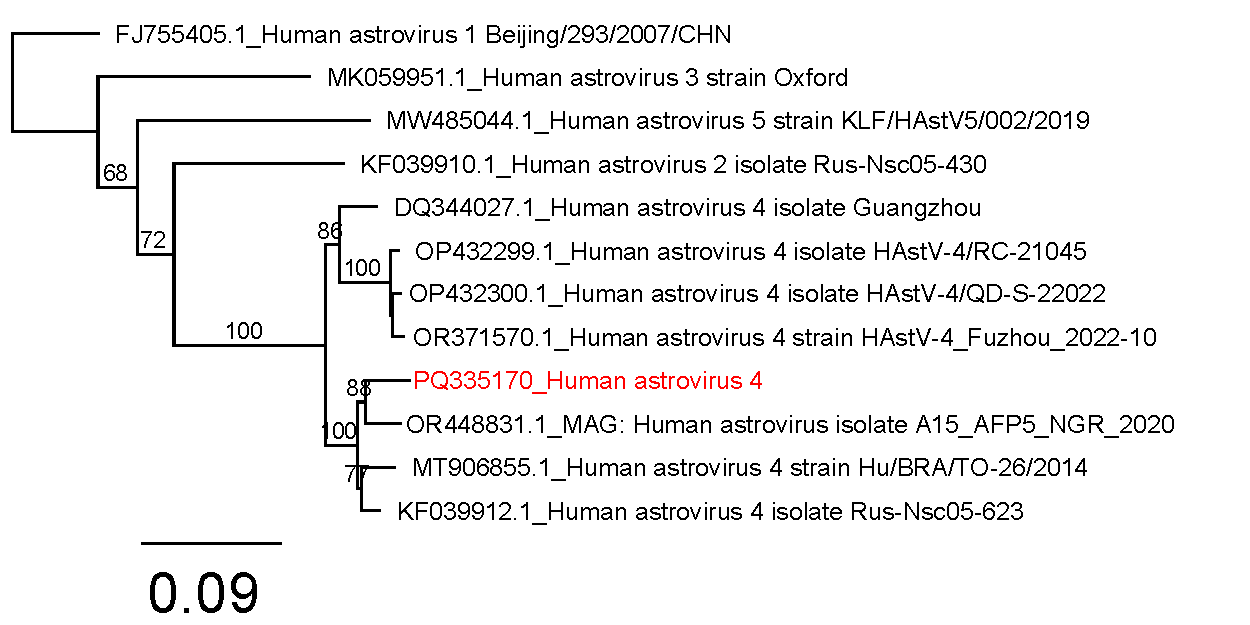

*Human astrovirus 4 (Mamastrovirus1)*

*Human polyomavirus 10* (MW polyomavirus), Deltapolyomavirus decihominis

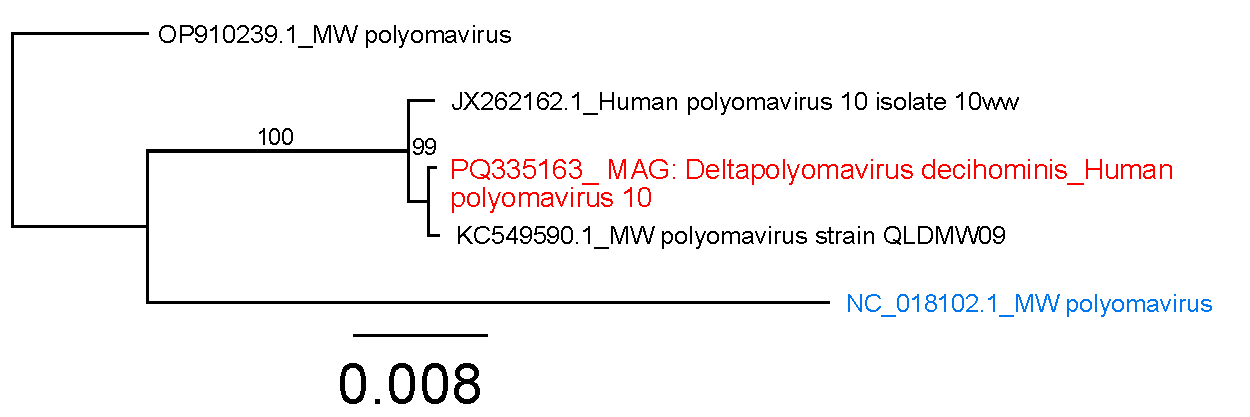

*WU polyomavirus – Betapolyomavirus quartihominis*

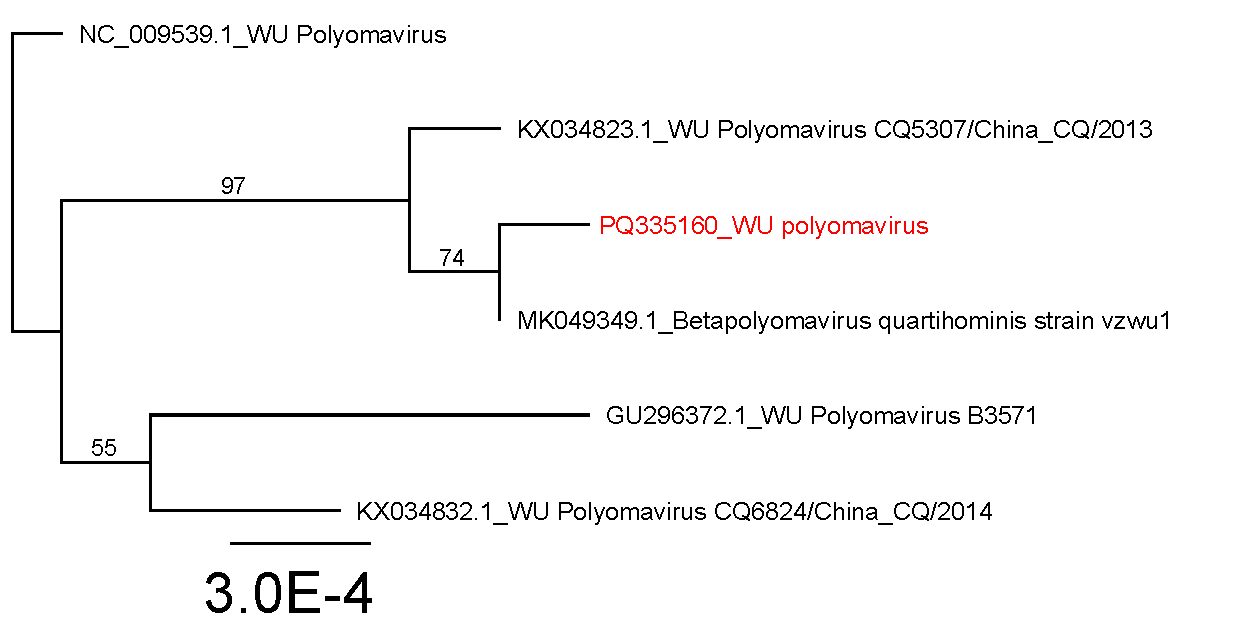

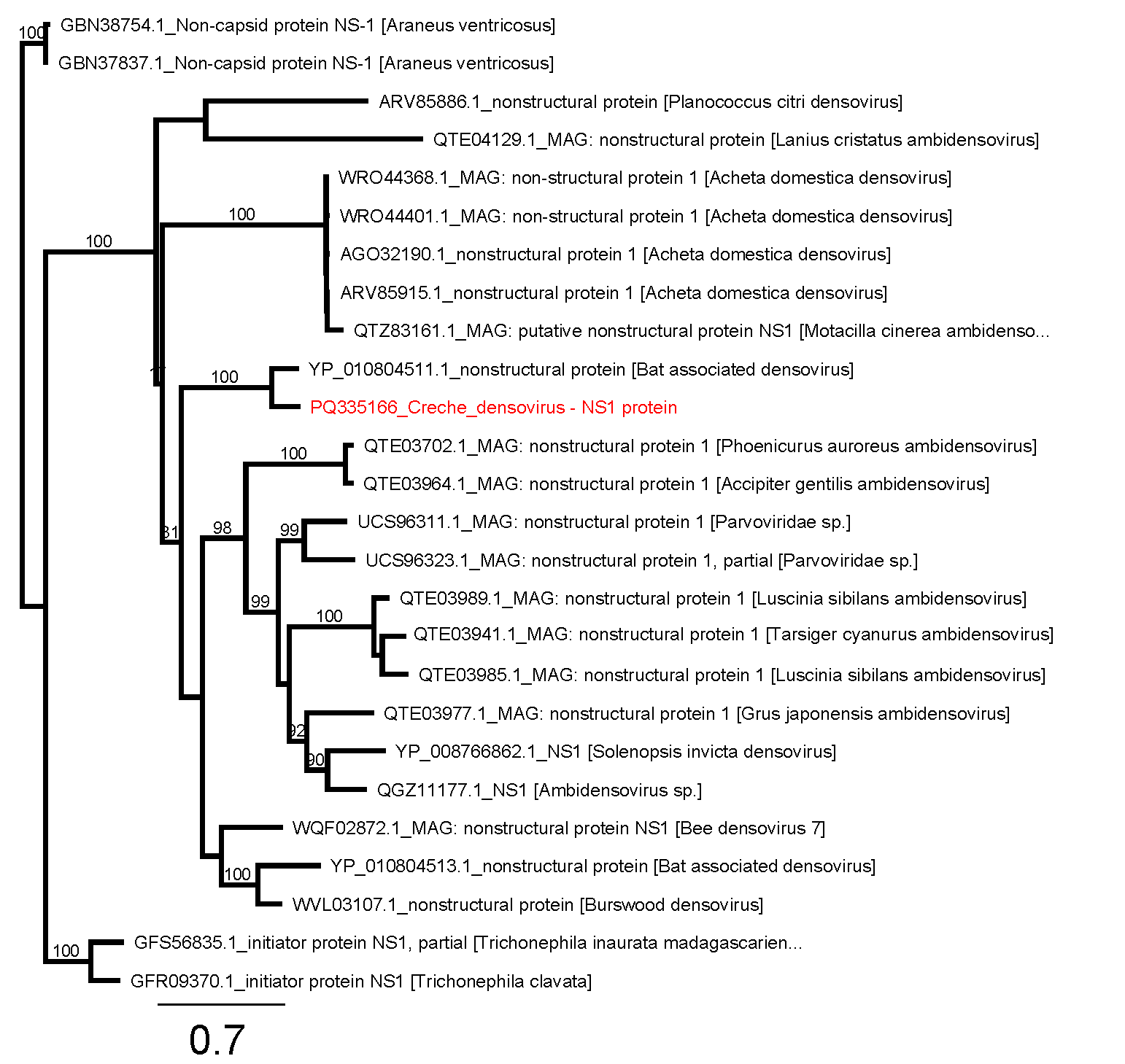
Red colored text shows the identified sequence from indoor air samples while blue-color sequences were recovered from the RefSeq database. All other sequences in black are sequences were sequences recovered from GenBank database. Phylogenetic trees were constructed using near-complete genomes (>95% horizontal coverage) for AAV-2 and human polyomaviruses. For human astrovirus 4, multiple contigs from ORF1a, ORF1b, and ORF2, comprising up to 71% of the genome (3.6 kbp), were combined to construct the phylogenetic tree. The phylogenetic tree for Canary polyomavirus was built using two contigs (1.9 kbp and 580 bp), covering both VP2 and VP1. The phylogenetic tree for Felis domesticus papillomavirus was constructed using a 4.4 kbp consensus sequence, corresponding to the E2 and L1 proteins. Phylogenetic tree of novel densovirus (*Creche densovirus*) was constructed using amino acid sequence of NS1 protein.

**Supplementary information 2. S3.** WU Polyomavirus number of differences among (near) complete genomes.

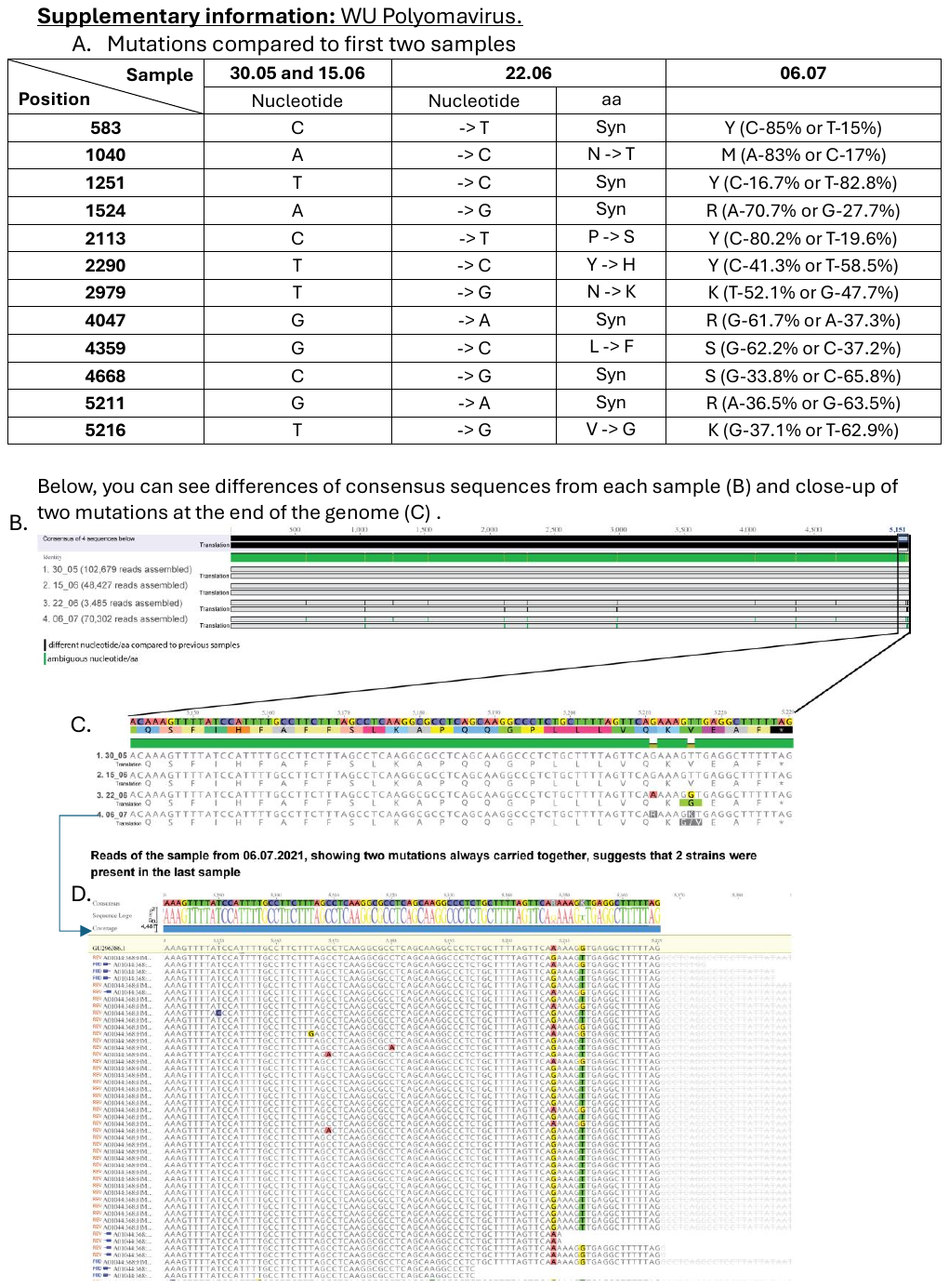

**Supplementary information 2. S4.** Metadata.

|  | sampling_Date | humidity_mean | temperature_mean | co2_mean | co2_max | sampling_duration | Month | Windows | #people |
| --- | --- | --- | --- | --- | --- | --- | --- | --- | --- |
| 1 | **10/01/2022** | 30.77 | 20.63 | 835.54 | 1000.00 | 2.25 | January | Closed | 15 |
| 2 | **19/01/2022** | 30.77 | 21.00 | 574.54 | 607.00 | 2.37 | January | Closed | 6 |
| 3 | **24/01/2022** | 29.10 | 20.40 | 594.90 | 717.00 | 1.88 | January | Closed | 10 |
| 4 | **26/01/2022** | 29.27 | 20.99 | 697.18 | 717.00 | 1.88 | January | Closed | 10 |
| 5 | **31/01/2022** | 34.82 | 20.31 | 829.27 | 917.00 | 1.95 | January | Closed | 15 |
| 6 | **07/02/2022** | 36.43 | 20.69 | 1009.00 | 1122.00 | 1.85 | February | Closed | 15 |
| 7 | **21/02/2022** | 32.55 | 20.46 | 827.64 | 1064.00 | 2.20 | February | Closed | 20 |
| 8 | **23/02/2022** | 34.23 | 21.29 | 709.77 | 868.00 | 2.08 | February | Closed | 16 |
| 9 | **04/03/2022** | 25.15 | 21.24 | 832.15 | 941.00 | 2.25 | March | Closed | 17 |
| 10 | **07/03/2022** | 24.64 | 21.69 | 1035.91 | 1119.00 | 2.03 | March | Closed | 23 |
| 11 | **09/03/2022** | 23.15 | 22.02 | 731.46 | 971.00 | 2.20 | March | Closed | 18 |
| 12 | **11/03/2022** | 24.43 | 21.86 | 711.21 | 776.00 | 2.25 | March | Closed | 23 |
| 13 | **14/03/2022** | 38.09 | 21.18 | 900.36 | 991.00 | 2.18 | March | Closed | 21 |
| 14 | **16/03/2022** | 34.09 | 21.83 | 820.73 | 894.00 | 1.95 | March | Closed | 20 |
| 15 | **21/03/2022** | 32.33 | 21.25 | 919.67 | 1165.00 | 2.25 | March | Closed | 22 |
| 16 | **23/03/2022** | 29.82 | 21.57 | 734.18 | 963.00 | 1.83 | March | Open | 18 |
| 17 | **30/03/2022** | 36.42 | 21.07 | 900.92 | 1161.00 | 2.42 | March | Closed | 17 |
| 18 | **04/04/2022** | 28.89 | 20.30 | 831.67 | 989.00 | 1.80 | April | Closed | 15 |
| 19 | **08/04/2022** | 35.45 | 20.81 | 910.09 | 1017.00 | 1.83 | April | Closed | 16 |
| 20 | **25/04/2022** | 38.70 | 21.35 | 925.00 | 985.00 | 2.05 | April | Open | 22 |
| 21 | **06/05/2022** | NA | NA | NA | NA | NA | May | Open | 20 |
| 22 | **11/05/2022** | NA | NA | NA | NA | NA | May | Open | 20 |
| 23 | **18/05/2022** | NA | NA | NA | NA | NA | May | Open | 16 |
| 24 | **23/05/2022** | NA | NA | NA | NA | NA | May | Open | 21 |
| 25 | **30/05/2022** | 40.20 | 21.22 | 730.30 | 826.00 | 2.13 | May | Open | 18 |
| 26 | **08/06/2022** | NA | NA | NA | NA | NA | June | Open | 18 |
| 27 | **15/06/2022** | NA | NA | NA | NA | NA | June | Open | 14 |
| 28 | **22/06/2022** | NA | NA | NA | NA | NA | June | Open | 19 |
| 29 | **29/06/2022** | 54.50 | 23.12 | 745.80 | 1030.00 | 1.92 | June | Open | 23 |
| 30 | **06/07/2022** | 47.94 | 22.48 | 779.72 | 1102.00 | 3.35 | July | Open | 18 |
| 31 | **13/07/2022** | 51.00 | 24.38 | 630.18 | 718.00 | 1.87 | July | Open | 18 |
| 32 | **07/09/2022** | NA | NA | NA | NA | NA | September | Open | 19 |
| 33 | **14/09/2022** | 58.17 | 23.19 | 636.17 | 866.00 | 2.02 | September | Open | 18 |
| 34 | **28/09/2022** | NA | NA | NA | NA | NA | September | Open | 20 |
| 35 | **05/10/2022** | NA | NA | NA | NA | NA | October | Open | NA |
| 36 | **12/10/2022** | 37.17 | 21.24 | 744.42 | 856.00 | 2.17 | October | Open | 20 |
| 37 | **19/10/2022** | 48.09 | 21.51 | 802.45 | 906.00 | 1.92 | October | Open | NA |
| 38 | **26/10/2022** | 54.42 | 21.93 | 798.58 | 912.00 | 2.08 | October | Open | 23 |
| 39 | **09/11/2022** | 48.00 | 21.69 | 704.29 | 780.00 | 2.07 | November | Closed | NA |
| 40 | **16/11/2022** | 47.00 | 21.70 | 924.30 | 1073.00 | 2.07 | November | Closed | NA |
| 41 | **23/11/2022** | NA | NA | NA | NA | NA | November | Closed | NA |
| 42 | **07/12/2022** | 36.73 | 21.06 | 929.64 | 1027.00 | 2.12 | December | Closed | NA |
